# Impact of Sepsis Identification and Treatment Time on In-Hospital Mortality, Length of Stay, and Hours in the Intensive Care Unit

**DOI:** 10.1101/2021.05.26.21257894

**Authors:** Sylvia EK Sudat

## Abstract

**Objective:** The objective of this study was to examine the impact of timely treatment and identification of sepsis on patient outcomes at Sutter Health, a mixed-payer healthcare system in northern California, US.

**Methods:** This observational, retrospective analysis considered electronic health record (EHR) data for individuals who presented with sepsis during 2016-17 at any of Sutter Health’s 22 emergency departments (ED). Impacts were assessed for the timing of broad-spectrum antibiotic and intravenous (IV) fluid initiation, first vital signs, sepsis screening, and lactate results. Outcomes were in-hospital mortality, hospital length of stay (LOS) and intensive care unit (ICU) hours for patients discharged alive.

**Results:** The final sample size was 35,847 (N=9,638 severe sepsis, N=5,309 septic shock). Early fluid initiation had the largest estimated impacts: a mortality reduction of 2.85%[2.03%,3.68%] overall and 2.94%[1.44%,4.48%] for severe sepsis (within 1 hour of sepsis presentation), and 14.66%[9.23%,20.07%] for septic shock (within 3 hours); reduced LOS (days) 1.39[1.08,1.71] overall, 2.30[1.31,3.21] severe sepsis, 3.07[1.21,4.94] septic shock; and fewer ICU hours 25.93[16.95,34.66] overall, 35.06[14.7,56.99] severe sepsis, 41.99[15.70,70.68] septic shock (within 3 hours). Sepsis screening within 30 minutes was also associated with mortality reductions (3.88%[2.96%,4.90%] overall, 1.74%[0.08%,3.50%] severe sepsis, 6.78%[3.12%,10.33%] septic shock). The greatest improvement opportunity was estimated for joint initiation of antibiotics and IV fluids, with a modest additional mortality reduction of 0.80%[0.47%,1.17%] overall, 0.77%[0.34%,1.19%] severe sepsis, 2.94%[1.83%,3.97%] septic shock; LOS reduction of 0.37[0.28,0.46] overall, 0.29[0.17,0.43] severe sepsis, 0.25[0.01,0.51] septic shock (within 1 hour); ICU hours reduction of 4.85[3.26,6.57] overall, 5.07[2.55,7.67] severe sepsis, 3.85[1.69,6.24] septic shock (within 3 hours).

## INTRODUCTION

Sepsis is a leading cause of in-hospital deaths, and early treatment of sepsis has been recognized as essential in the promotion of better outcomes.^1,2^ Septic patients on average also stay longer in the hospital, and the costs of sepsis management represent a substantial economic burden.^1,3^ The Surviving Sepsis Campaign (survivingsepsis.org) has provided guidance on implementation of 6-hour, 3-hour, and finally 1-hour bundles for sepsis care, and there is evidence that implementation of these guidelines has improved timeliness of sepsis treatment and outcomes for people with sepsis around the world.^4-25^ The Center for Medicare and Medicaid Services (CMS) in the US has also established a quality measure for hospitals based on compliance with a sepsis treatment bundle (SEP-1), although the bundle has received criticism, and there does not appear to be evidence that its implementation resulted in improved outcomes for people with sepsis.^26-33^

This study is a rigorous evaluation of sepsis treatment and identification timing on patient outcomes at 22 hospitals in a large health system during 2016-17. The specific focus is on adult patients whose sepsis onset occurred during or before their time in the emergency department (ED), and who went on to be admitted to the hospital.

Considered in this study were the timing of broad-spectrum antibiotic and intravenous (IV) fluid initiation, first vital signs, documentation of sepsis screening, and initial lactate laboratory test results. The following patient outcomes were measured: in-hospital mortality, hospital length of stay (LOS) for patients discharged alive, and hours spent in the intensive care unit (ICU) for patients who went to the ICU and were discharged alive. Impact was measured in two ways: (1) as the difference between 0% success and 100% success, and (2) as the difference between current practice and 100% success. The second estimate represents the potential opportunity presented for making additional improvements, based on current (2016-17) success rates.

## METHODS

### Analytical population

Sutter Health is a large, mixed-payer healthcare system in northern California, US. Sutter serves approximately 3.5 million patients per year in both the acute (hospital) and non-acute (e.g. physician office) setting. The analytical population for this study comprises 2016-17 adult hospital admissions at any of Sutter Health’s 22 acute care hospitals with emergency departments meeting the following criteria: (1) admission began in the ED, (2) severe sepsis clinical criteria were met (according to vital signs and lab values, see next section) while in the ED, and (3) treatment with broad-spectrum antibiotics and intravenous (IV) fluids (any volume) was initiated at any point during hospitalization (either while in the ED or after inpatient admission). The following were excluded: transfers from other healthcare facilities/hospitals, admissions for labor and delivery, elective admissions, and non-acute admissions (behavioral health, rehabilitation, and hospice).

### Severe sepsis clinical criteria

We followed the CMS definition of severe sepsis – at least two symptoms of systemic inflammatory response syndrome (SIRS), and one symptom of organ dysfunction (see below).^26^ The definition of sepsis has changed over time; the newest definition no longer relies upon SIRS criteria, and the designation “ severe sepsis” has been eliminated.^34^ However, the older definition is used here because it most closely aligns with hospital definitions of sepsis at the time of this study (2016-17).

For the purposes of this study, the term “ sepsis onset” will refer to the first time these clinical criteria were met according to data in the medical record. This has also been referred to as sepsis “ time zero.”

SIRS criteria:

- Temperature > 38.3°C or < 36.0°C
- Heart rate > 90
- Respiratory rate > 20 breaths/min or pC0_2_< 32
- White blood cell count > 12,000 or < 4,000 or > 10% bands

Signs of organ dysfunction:

- Systolic blood pressure (SBP) < 90
- Mean arterial pressure (MAP) < 65
- Lactate > 2 mmol/L
- Creatinine > 2 mg/dL
- Bilirubin > 2 mg/ddL
- Platelets < 100,000
- INR > 1.5 or aPTT > 60 sec
- Altered mental state (Glasgow coma scale < 15)

### qSOFA criteria

Patients are at high risk of adverse outcomes, according to the quick Sequential [Sepsis-related] Organ Failure Assessment (qSOFA),^34,35^ if they meet any two of the following clinical criteria:

- Altered mental state
- Respiratory rate 22 breaths/min or more
- SBP 100 mm Hg or less

The qSOFA was used in statistical models for severity adjustment, as described below.

### Statistical methods

The impact of timely treatment was assessed by considering initiation of broad-spectrum antibiotics and IV fluids, both singly and in combination, within 1 and 3 hours of ED arrival and of sepsis onset. Because the focus of this study was the timing of treatment, no restriction was placed on the volume of IV fluids given, only on the timing of fluid initiation. To address the impact of early sepsis identification on outcomes, the following were considered, measured from ED arrival: vital signs taken within 10 minutes, sepsis screening documented within 15 or 30 minutes, and lactate resulted within 1 hour or 3 hours. The patient outcomes considered were in-hospital mortality, hospital LOS for patients discharged alive, and hours spent in the ICU for patients who went to the ICU and were discharged alive.

To assess impact, a counterfactual approach was used.^36^ Specifically, each outcome was modeled separately, and used to predict the expected result if the treatment or identification event was 100% successful or 0% successful – for example, if all patients were treated with broad-spectrum antibiotics within 1 hour of sepsis onset, versus being treated after 1 hour (G-computation estimator).^37^ We used logistic regression to model mortality, and negative binomial regression for hospital LOS and hours in the ICU. Separate models (153 in total) were constructed for each treatment/identification variable, outcome, and cohort (full cohort, severe sepsis, and septic shock).

We included the following variables in the outcome models estimating projected outcomes: patient demographics (age category by decade from 60-80+, sex, race, ethnicity), clinical characteristics (comorbidities, Charlson comorbidity index), prior utilization (number of hospitalizations in the prior year, number of ED visits in the prior 6 months), information on prior infection (sepsis diagnosis in the prior year, number of cellulitis diagnoses in the prior year, number of urinary tract infections in the prior year), proxies for severity at presentation (sepsis clinical criteria met at sepsis onset, qSOFA at sepsis onset, time to sepsis onset), and the probability of developing severe sepsis or septic shock based on clinical criteria at sepsis presentation.^38-40^ Time to sepsis onset was included as a severity measure because patients with early onset were observed to have higher mortality (see Appendix). The probability of developing severe sepsis or septic shock was modeled separately using logistic regression. The lactate criterion was excluded from the 18 models estimating impact of time to lactate result.

In each case, we computed two estimates of impact: (1) the difference between 0% success and 100% success, and (2) the difference between current practice and 100% success. Confidence intervals for impact estimates were constructed using the nonparametric bootstrap with 1000 iterations.

## RESULTS AND DISCUSSION

The study population is described in Figure 1. Out of a total of 357,012 adult hospital admissions to Sutter Health acute care hospitals in 2016 and 2017, 41,757 (12%) were associated with a sepsis discharge diagnosis unrelated to childbirth or miscarriage. Of these, 253,331 (71%) met basic inclusion criteria (after applying exclusions described in the Methods section), including 39,210 (94%) of sepsis admissions. Severe sepsis clinical criteria were met while in the ED for 51,701 (20%) of all admissions and 20,736 (53%) of sepsis admissions meeting basic inclusion criteria. Finally, 35,847 (69%) of these patients were also treated with broad-spectrum antibiotics and IV fluids during hospitalization, including 19,675 (95%) with sepsis discharge diagnosis. The final study population therefore represents 10% of all 2016-17 adult hospital admissions and 47% of sepsis admissions.

**Figure 1.**
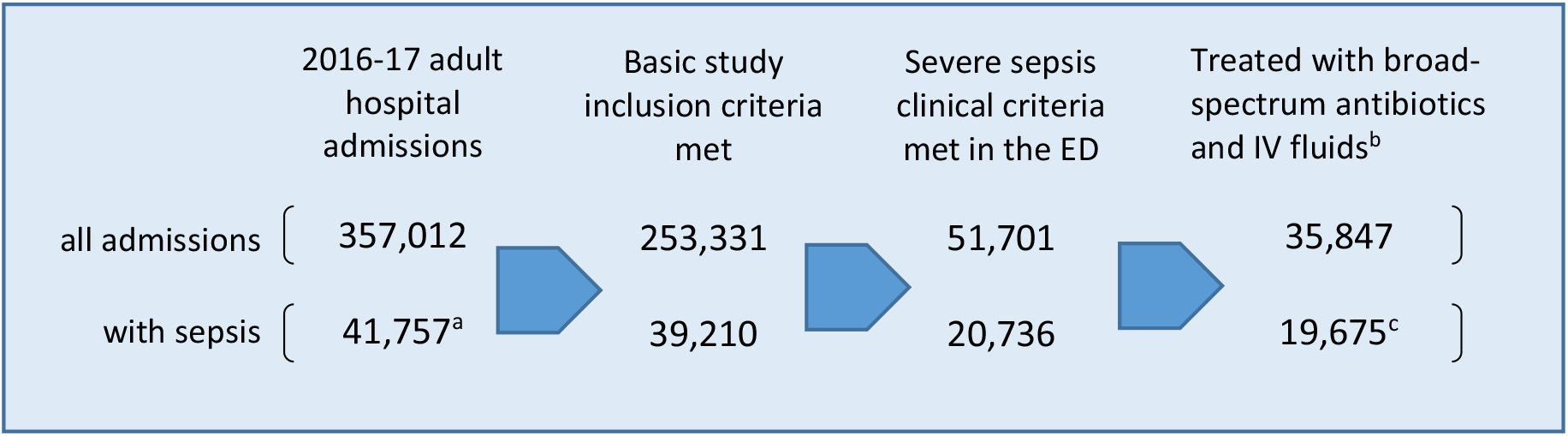
Construction of study population ^a^14,581 simple, 18,052 severe, 9,124 shock (discharge diagnosis); ^b^Treatment with IV fluids (any volume) and broad-spectrum antibiotics at any point during hospitalization, not necessarily in the ED; ^c^4,728 simple, 9,638 severe, 5,309

The overall mortality rate for the study population was 8.58% (5.89% severe sepsis, 24.68% septic shock). Patients were discharged alive for 32,771 admissions (9,068 severe sepsis, 3,999 septic shock), and the average hospital LOS was 6.68 days overall, 6.62 for severe sepsis, and 10.12 for septic shock. ICU stays were part of 10,544 admissions where patients were discharged alive, including 2,395 of those for severe sepsis and 3,488 of those for septic shock. Patients spent on average 89.43 hours in the ICU (76.24 severe sepsis, 109.93 septic shock). Patients were screened for sepsis 99% of the time (N=35,837), and had a documented lactate lab result 93% of the time (N=33,472).

Descriptive data for the N=35,847 full study population are shown in Table 1. Data are summarized by admission, not by unique patient. The cohort was 63.8% white, 11.7% Black/African American, 11.1% Asian, and 2.54% other races. For 10.9% of admissions patient race was unknown or not reported. Hispanic ethnicity was reported for 15.4% of admissions. The most common comorbidities were diabetes without chronic complications (24.4%), congestive heart failure (19.1%), chronic pulmonary disease (25.5%), and renal disease (22.5%). Just over half of admissions were associated with a sepsis discharge diagnosis (13.2% simple, 26.9% severe, 14.8% shock), and 45.1% had no sepsis diagnosis. On average, broad-spectrum antibiotics were administered 2.5 hours (153 minutes) after ED arrival, and just under an hour (58 minutes) after severe sepsis clinical criteria were met (sepsis onset). IV fluids were first given about an hour (66 minutes) after ED arrival, and 5 minutes prior to sepsis onset. The median time to lactate result was 1.5 hours (79 minutes) after arrival in the ED. On average, vital signs were first taken on 5 minutes after arrival, and sepsis screening was documented 8 minutes after arrival.

**Table 1.**
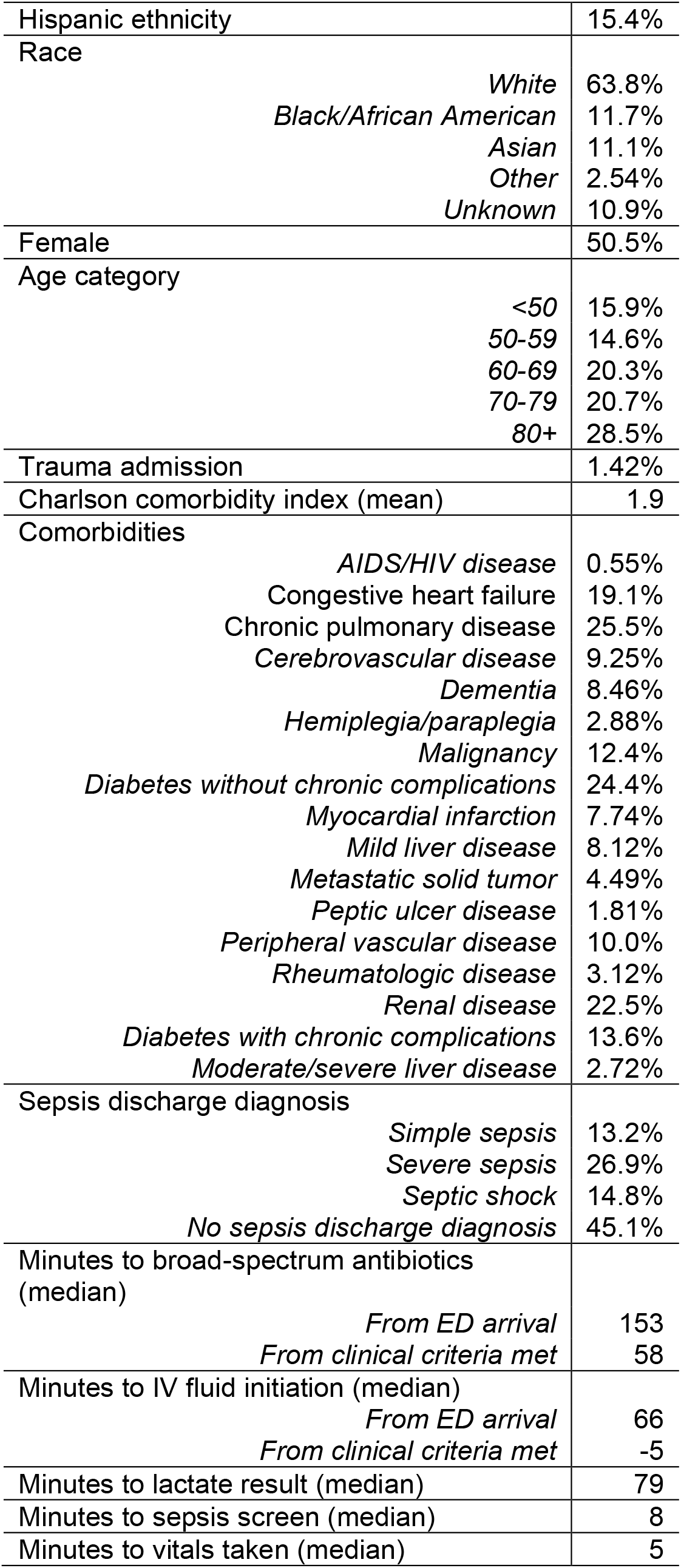
Study population descriptive data (N=35,847)

|  |  |
| --- | --- |
| Hispanic ethnicity | 15.4% |
| Race |  |
| *White* | 63.8% |
| *Black/African American* | 11.7% |
| *Asian* | 11.1% |
| *Other* | 2.54% |
| *Unknown* | 10.9% |
| Female | 50.5% |
| Age category |  |
| <50 | 15.9% |
| 50-59 | 14.6% |
| 60-69 | 20.3% |
| 70-79 | 20.7% |
| 80+ | 28.5% |
| Trauma admission | 1.42% |
| Charlson comorbidity index (mean) | 1.9 |
| Comorbidities |  |
| *AIDS/HIV disease* | 0.55% |
| Congestive heart failure | 19.1% |
| Chronic pulmonary disease | 25.5% |
| *Cerebrovascular disease* | 9.25% |
| *Dementia* | 8.46% |
| *Hemiplegia/paraplegia* | 2.88% |
| *Malignancy* | 12.4% |
| *Diabetes without chronic complications* | 24.4% |
| *Myocardial infarction* | 7.74% |
| *Mild liver disease* | 8.12% |
| *Metastatic solid tumor* | 4.49% |
| *Peptic ulcer disease* | 1.81% |
| *Peripheral vascular disease* | 10.0% |
| *Rheumatologic disease* | 3.12% |
| *Renal disease* | 22.5% |
| *Diabetes with chronic complications* | 13.6% |
| *Moderate/severe liver disease* | 2.72% |
| Sepsis discharge diagnosis |  |
| *Simple sepsis* | 13.2% |
| *Severe sepsis* | 26.9% |
| *Septic shock* | 14.8% |
| *No sepsis discharge diagnosis* | 45.1% |
| Minutes to broad-spectrum antibiotics (median) |  |
| *From ED arrival* | 153 |
| *From clinical criteria met* | 58 |
| Minutes to IV fluid initiation (median) |  |
| *From ED arrival* | 66 |
| *From clinical criteria met* | -5 |
| Minutes to lactate result (median) | 79 |
| Minutes to sepsis screen (median) | 8 |
| Minutes to vitals taken (median) | 5 |

Tables 2-4 display the estimated impact of treatment with broad-spectrum antibiotics and IV fluids within 1 and 3 hours of sepsis onset (defined as first emergence of severe sepsis clinical criteria in the medical record) on mortality, hospital LOS, and hours in the ICU. Tables 5-7 are analogous to Tables 2-4, but measure time-to-treatment from ED arrival instead of sepsis onset. Finally, Tables 8-10 show the impact on outcomes (mortality, hospital LOS, ICU hours) of the identification-related measures: time to lactate result, time to sepsis screen, and time to first vital sign taken.

**Table 2.** Estimated impact of timely treatment on in-hospital mortality, measured from first emergence of severe sepsis symptoms in the medical record.

|  |  |  | Expected mortality<br>by % treated |  | Expected impact of 100% treatment (95% CI) |  |
| --- | --- | --- | --- | --- | --- | --- |
| Treatment | Time<br>(from CC) | % treated | 0% treated | 100% treated | Difference: 0% vs. 100% | Difference: current<sup>a</sup> % treated<br>vs. 100% |
| Full cohort<sup>b</sup> (N=35,847). 2016-17 mortality: 8.58% |  |  |  |  |  |  |
| Broad-spectrum antibiotics | 1 hour | 51.2% | 9.07% | 8.06% | -1.01% (-1.65%,-0.40%)<sup>d</sup> | -0.51% (-0.84%,-0.21%)<sup>d</sup> |
| Broad-spectrum antibiotics | 3 hours | 74.6% | 9.92% | 8.11% | -1.80% (-2.56%,-1.12%)<sup>d</sup> | -0.46% (-0.65%,-0.29%)<sup>d</sup> |
| IV fluids<sup>c</sup> | 1 hour | 81.1% | 10.87% | 8.03% | -2.85% (-3.68%,-2.03%)<sup>d</sup> | -0.55% (-0.71%,-0.39%)<sup>d</sup> |
| IV fluids | 3 hours | 89.8% | 11.08% | 8.30% | -2.79% (-3.86%,-1.68%)<sup>d</sup> | -0.28% (-0.39%,-0.17%)<sup>d</sup> |
| Broad-spectrum antibiotics<br>+ IV fluids | 1 hour | 46.6% | 29.23% | 7.77% | -1.46% (-2.14%,-0.85%)<sup>d</sup> | -0.80% (-1.17%,-0.47%)<sup>d</sup> |
| Broad-spectrum antibiotics<br>+ IV fluids | 3 hours | 69.8% | 9.97% | 7.98% | -1.99% (-2.68%,-1.33%)<sup>d</sup> | -0.60% (-0.80%,-0.40%)<sup>d</sup> |
| Severe sepsis (N= 9,638). 2016-17 mortality: 5.89% |  |  |  |  |  |  |
| Broad-spectrum antibiotics | 1 hour | 61.5% | 6.50% | 5.47% | -1.03% (-2.00%,-0.04%)<sup>d</sup> | -0.42% (-0.82%,-0.01%)<sup>d</sup> |
| Broad-spectrum antibiotics | 3 hours | 86.2% | 6.61% | 5.78% | -0.83% (-2.31%,+0.52%) | -0.12% (-0.32%,+0.07%) |
| IV Fluids | 1 hour | 86.6% | 8.38% | 5.44% | -2.94% (-4.48%,-1.44%)<sup>d</sup> | -0.46% (-0.68%,-0.23%)<sup>d</sup> |
| IV Fluids | 3 hours | 94.2% | 8.41% | 5.72% | -2.70% (-4.96%,-0.57%)<sup>d</sup> | -0.18% (-0.33%,-0.04%)<sup>d</sup> |
| Broad-spectrum antibiotics<br>+ IV fluids | 1 hour | 57.3% | 6.83% | 5.12% | -1.71% (-2.65%,-0.73%)<sup>d</sup> | -0.77% (-1.19%,-0.34%)<sup>d</sup> |
| Broad-spectrum antibiotics<br>+ IV fluids | 3 hours | 82.4% | 7.08% | 5.63% | -1.45% (-2.78%,-0.13%)<sup>d</sup> | -0.26% (-0.49%,-0.02%)<sup>d</sup> |
| Septic shock (N= 5,309). 2016-17 mortality: 24.68% |  |  |  |  |  |  |
| Broad-spectrum antibiotics | 1 hour | 60.3% | 28.12% | 22.32% | -5.80% (-8.48%,-3.43%)<sup>d</sup> | -2.36% (-3.42%,-1.40%)<sup>d</sup> |
| Broad-spectrum antibiotics | 3 hours | 85.6% | 33.02% | 23.24% | -9.79% (-13.48%,-6.01%)<sup>d</sup> | -1.44% (-2.00%,-0.89%)<sup>d</sup> |
| IV fluids | 1 hour | 87.1% | 33.00% | 23.37% | -9.64% (-13.13%,-6.09%)<sup>d</sup> | -1.31% (-1.78%,-0.83%)<sup>d</sup> |
| IV fluids | 3 hours | 94.9% | 38.56% | 23.89% | -14.66% (-20.07%,-9.23%)<sup>d</sup> | -0.78% (-1.09%,-0.49%)<sup>d</sup> |
| Broad-spectrum antibiotics<br>+ IV fluids | 1 hour | 56.7% | 28.37% | 21.74% | -6.63% (-9.05%,-4.19%)<sup>d</sup> | -2.94% (-3.97%,-1.83%)<sup>d</sup> |
| Broad-spectrum antibiotics<br>+ IV fluids | 3 hours | 83.0% | 33.56% | 22.81% | -10.75% (-14.36%,-7.28%)<sup>d</sup> | -1.86% (-2.46%,-1.26%)<sup>d</sup> |

CC=severe sepsis clinical criteria met; CI=confidence interval; ED=emergency department; IV=intravenous
<sup>a</sup>2016-2017; <sup>b</sup>Any admission where (1) severe sepsis clinical criteria were met while in the ED, and (2) patient was treated with broad-spectrum antibiotics and IV fluids (any volume) at some point during hospitalization; <sup>c</sup>Any volume; <sup>d</sup>Statistically significant (p<0.05)

**Table 3.** Estimated impact of timely treatment on mean hospital LOS (days) for patients discharged alive, measured from first emergence of severe sepsis symptoms in the medical record.

|  |  |  | Expected mean hospital LOS<br>by % treated |  | Expected impact of 100% treatment (95% CI) |  |
| --- | --- | --- | --- | --- | --- | --- |
| Treatment | Time<br>(from CC) | % treated | 0% treated | 100% treated | Difference: 0% vs. 100% | Difference: current<sup>a</sup> % treated vs.<br>100% |
| Full cohort<sup>b</sup> (N= 32,771). 2016-17 mean hospital LOS: 6.68 days |  |  |  |  |  |  |
| Broad-spectrum antibiotics | 1 hour | 51.6% | 6.96 | 6.39 | -0.57 (-0.73,-0.41)<sup>d</sup> | -0.28 (-0.36,-0.20)<sup>d</sup> |
| Broad-spectrum antibiotics | 3 hours | 75.0% | 7.44 | 6.40 | -1.04 (-1.25,-0.83)<sup>d</sup> | -0.27 (-0.33,-0.22)<sup>d</sup> |
| IV fluids<sup>c</sup> | 1 hour | 81.5% | 7.47 | 6.49 | -0.98 (-1.22,-0.74)<sup>d</sup> | -0.19 (-0.23,-0.14)<sup>d</sup> |
| IV fluids | 3 hours | 90.1% | 7.92 | 6.53 | -1.39 (-1.71,-1.08)<sup>d</sup> | -0.14 (-0.18,-0.11)<sup>d</sup> |
| Broad-spectrum antibiotics<br>+ IV fluids | 1 hour | 47.1% | 6.99 | 6.30 | -0.69 (-0.85,-0.53)<sup>d</sup> | -0.37 (-0.46,-0.28)<sup>d</sup> |
| Broad-spectrum antibiotics<br>+ IV fluids | 3 hours | 70.2% | 7.45 | 6.33 | -1.12 (-1.33,-0.92)<sup>d</sup> | -0.35 (-0.41,-0.28)<sup>d</sup> |
| Severe sepsis (N= 9,068). 2016-17 mean hospital LOS: 6.62 days |  |  |  |  |  |  |
| Broad-spectrum antibiotics | 1 hour | 61.9% | 6.91 | 6.42 | -0.50 (-0.83,-0.18)<sup>d</sup> | -0.20 (-0.33,-0.08)<sup>d</sup> |
| Broad-spectrum antibiotics | 3 hours | 86.3% | 7.66 | 6.43 | -1.23 (-1.78,-0.74)<sup>d</sup> | -0.19 (-0.27,-0.12)<sup>d</sup> |
| IV Fluids | 1 hour | 87.1% | 7.74 | 6.43 | -1.31 (-1.98,-0.70)<sup>d</sup> | -0.18 (-0.28,-0.10)<sup>d</sup> |
| IV Fluids | 3 hours | 94.4% | 8.77 | 6.47 | -2.30 (-3.21,-1.31)<sup>d</sup> | -0.14 (-0.21,-0.08)<sup>d</sup> |
| Broad-spectrum antibiotics<br>+ IV fluids | 1 hour | 57.9% | 6.99 | 6.32 | -0.67 (-0.97,-0.37)<sup>d</sup> | -0.29 (-0.43,-0.17)<sup>d</sup> |
| Broad-spectrum antibiotics<br>+ IV fluids | 3 hours | 82.7% | 7.72 | 6.36 | -1.36 (-1.82,-0.93)<sup>d</sup> | -0.25 (-0.35,-0.17)<sup>d</sup> |
| Septic shock (N= 3,999). 2016-17 mean hospital LOS: 10.12 days |  |  |  |  |  |  |
| Broad-spectrum antibiotics | 1 hour | 62.5% | 10.26 | 10.02 | -0.24 (-0.88,+0.35) | -0.10 (-0.34,+0.12) |
| Broad-spectrum antibiotics | 3 hours | 87.4% | 11.27 | 9.93 | -1.34 (-2.30,-0.40)<sup>d</sup> | -0.19 (-0.32,-0.06)<sup>d</sup> |
| IV fluids | 1 hour | 88.9% | 11.47 | 9.94 | -1.53 (-2.56,-0.51)<sup>d</sup> | -0.19 (-0.32,-0.07)<sup>d</sup> |
| IV fluids | 3 hours | 96.0% | 13.05 | 9.98 | -3.07 (-4.94,-1.21)<sup>d</sup> | -0.14 (-0.24,-0.06)<sup>d</sup> |
| Broad-spectrum antibiotics<br>+ IV fluids | 1 hour | 59.3% | 10.45 | 9.87 | -0.58 (-1.18,+0.01) | -0.25 (-0.51,-0.01)<sup>d</sup> |
| Broad-spectrum antibiotics<br>+ IV fluids | 3 hours | 85.2% | 11.52 | 9.85 | -1.67 (-2.58,-0.72)<sup>d</sup> | -0.27 (-0.43,-0.12)<sup>d</sup> |

CC=severe sepsis clinical criteria met; CI=confidence interval; ED=emergency department; IV=intravenous; LOS=length of stay
<sup>a</sup>2016-2017; <sup>b</sup>Any admission where (1) severe sepsis clinical criteria were met while in the ED, and (2) patient was treated with broad-spectrum antibiotics and IV fluids (any volume) at some point during hospitalization; <sup>c</sup>Any volume; <sup>d</sup>Statistically significant (p<0.05)

**Table 4.** Estimated impact of timely treatment on hours in the ICU, measured from first emergence of severe sepsis symptoms in the medical record.

|  |  |  | Expected mean ICU hours<br>by % treated |  | Expected impact of 100% treatment (95% CI) |  |
| --- | --- | --- | --- | --- | --- | --- |
| Treatment | Time<br>(from CC) | % treated | 0% treated | 100% treated | Difference: 0% vs. 100% | Difference: current<sup>a</sup> % treated vs.<br>100% |
| Full cohort<sup>b</sup> (N=10,544). 2016-17 mean time in ICU: 89.43 hours |  |  |  |  |  |  |
| Broad-spectrum antibiotics | 1 hour | 50.8% | 91.34 | 87.42 | -3.93 (-8.83,+0.10) | -2.01 (-4.43,+0.01) |
| Broad-spectrum antibiotics | 3 hours | 73.8% | 97.67 | 86.24 | -11.43 (-17.04,-6.83)<sup>d</sup> | -3.19 (-4.73,-1.96)<sup>d</sup> |
| IV fluids<sup>c</sup> | 1 hour | 81.7% | 106.32 | 85.49 | -20.83 (-27.24,-14.57)<sup>d</sup> | -3.94 (-5.18,-2.78)<sup>d</sup> |
| IV fluids | 3 hours | 90.2% | 112.7 | 86.77 | -25.93 (-34.66,-16.95)<sup>d</sup> | -2.66 (-3.57,-1.78)<sup>d</sup> |
| Broad-spectrum antibiotics<br>+ IV fluids | 1 hour | 46.4% | 92.80 | 85.27 | -7.53 (-12.43,-3.29)<sup>d</sup> | -4.16 (-6.87,-1.84)<sup>d</sup> |
| Broad-spectrum antibiotics<br>+ IV fluids | 3 hours | 69.5% | 99.72 | 84.58 | -15.14 (-20.64,-10.13)<sup>d</sup> | -4.85 (-6.57,-3.26)<sup>d</sup> |
| Severe sepsis (N=2,395). 2016-17 mean time in ICU: 76.24 hours |  |  |  |  |  |  |
| Broad-spectrum antibiotics | 1 hour | 61.3% | 76.92 | 74.9 | -2.02 (-11.25,+5.33) | -1.34 (-4.87,+1.51) |
| Broad-spectrum antibiotics | 3 hours | 84.6% | 86.11 | 73.67 | -12.44 (-24.75,-1.89)<sup>d</sup> | -2.57 (-4.97,-0.68)<sup>d</sup> |
| IV Fluids | 1 hour | 84.8% | 90.83 | 72.81 | -18.01 (-29.87,-6.27)<sup>d</sup> | -3.42 (-5.73,-1.4)<sup>d</sup> |
| IV Fluids | 3 hours | 92.2% | 107.88 | 72.81 | -35.06 (-56.99,-14.7)<sup>d</sup> | -3.42 (-5.62,-1.52)<sup>d</sup> |
| Broad-spectrum antibiotics<br>+ IV fluids | 1 hour | 55.9% | 80.32 | 71.96 | -8.36 (-16.89,-0.92)<sup>d</sup> | -4.28 (-8.02,-0.78)<sup>d</sup> |
| Broad-spectrum antibiotics<br>+ IV fluids | 3 hours | 79.6% | 92.65 | 71.17 | -21.48 (-32.86,-10.37)<sup>d</sup> | -5.07 (-7.67,-2.55)<sup>d</sup> |
| Septic shock (N=3,488). 2016-17 mean time in ICU: 109.93 hours |  |  |  |  |  |  |
| Broad-spectrum antibiotics | 1 hour | 62.8% | 109.84 | 109.71 | -0.13 (-8.90,+8.19) | -0.22 (-3.74,+3.05) |
| Broad-spectrum antibiotics | 3 hours | 87.5% | 121.4 | 107.95 | -13.44 (-27.08,-0.58)<sup>d</sup> | -1.98 (-3.92,-0.29)<sup>d</sup> |
| IV fluids | 1 hour | 88.7% | 128.16 | 107.3 | -20.86 (-37.00,-5.88)<sup>d</sup> | -2.64 (-4.82,-0.82)<sup>d</sup> |
| IV fluids | 3 hours | 95.9% | 149.9 | 107.91 | -41.99 (-70.68,-15.70)<sup>d</sup> | -2.02 (-3.53,-0.88)<sup>d</sup> |
| Broad-spectrum antibiotics<br>+ IV fluids | 1 hour | 59.4% | 112.6 | 107.71 | -4.89 (-13.61,+4.57) | -2.22 (-6.03,+1.67) |
| Broad-spectrum antibiotics<br>+ IV fluids | 3 hours | 85.3% | 129.58 | 106.08 | -23.5 (-37.36,-10.01)<sup>d</sup> | -3.85 (-6.24,-1.69)<sup>d</sup> |

CC=severe sepsis clinical criteria met; CI=confidence interval; ED=emergency department; ICU=intensive care unit; IV=intravenous
Analysis includes patients discharged alive who went to the ICU at some point during hospitalization.
<sup>a</sup>2016-2017; <sup>b</sup>Any admission where (1) severe sepsis clinical criteria were met while in the ED, and (2) patient was treated with broad-spectrum antibiotics and IV fluids (any volume) at some point during hospitalization; <sup>c</sup>Any volume; <sup>d</sup>Statistically significant (p<0.05)

**Table 5.** Estimated impact of timely treatment on in-hospital mortality, measured from ED arrival.

|  |  |  | Expected mortality<br>by % treated |  | Expected impact of 100% treatment (95% CI) |  |
| --- | --- | --- | --- | --- | --- | --- |
| Treatment | Time<br>(from ED-A) | % treated | 0% treated | 100% treated | Difference: 0% vs. 100% | Difference: current<sup>a</sup> % treated<br>vs. 100% |
| Full cohort<sup>b</sup> (N=35,847). 2016-17 mortality: 8.58% |  |  |  |  |  |  |
| Broad-spectrum antibiotics | 1 hour | 13.50% | 8.47% | 9.15% | +0.68% (-0.16%,+1.53%) | +0.57% (-0.13%,+1.28%) |
| Broad-spectrum antibiotics | 3 hours | 57.30% | 9.20% | 8.19% | -1.01% (-1.67%,-0.38%)<sup>d</sup> | -0.38% (-0.63%,-0.15%)<sup>d</sup> |
| IV fluids<sup>c</sup> | 1 hour | 46.70% | 8.58% | 8.57% | -0.01% (-0.60%,+0.62%) | -0.01% (-0.28%,+0.29%) |
| IV fluids | 3 hours | 81.20% | 10.65% | 8.20% | -2.46% (-3.37%,-1.56%)<sup>d</sup> | -0.38% (-0.52%,-0.24%)<sup>d</sup> |
| Broad-spectrum antibiotics<br>+ IV fluids | 1 hour | 11.80% | 8.52% | 8.91% | 0.39% (-0.53%,+1.27%) | +0.33% (-0.46%,+1.1%) |
| Broad-spectrum antibiotics<br>+ IV fluids | 3 hours | 52.80% | 9.25% | 8.08% | -1.17% (-1.82%,-0.6%)<sup>d</sup> | -0.49% (-0.76%,-0.25%)<sup>d</sup> |
| Severe sepsis (N= 9,638). 2016-17 mortality: 5.89% |  |  |  |  |  |  |
| Broad-spectrum antibiotics | 1 hour | 17.80% | 5.82% | 6.21% | 0.39% (-0.82%,+1.61%) | 0.31% (-0.66%,+1.30%) |
| Broad-spectrum antibiotics | 3 hours | 71.40% | 6.13% | 5.81% | -0.32% (-1.46%,+0.77%) | -0.08% (-0.39%,+0.20%) |
| IV Fluids | 1 hour | 51.20% | 6.20% | 5.62% | -0.58% (-1.54%,+0.44%) | -0.27% (-0.72%,+0.21%) |
| IV Fluids | 3 hours | 87.40% | 7.62% | 5.66% | -1.96% (-3.64%,-0.36%)<sup>d</sup> | -0.24% (-0.42%,-0.04%)<sup>d</sup> |
| Broad-spectrum antibiotics<br>+ IV fluids | 1 hour | 15.70% | 5.95% | 5.61% | -0.34% (-1.54%,+0.95%) | -0.28% (-1.28%,+0.79%) |
| Broad-spectrum antibiotics<br>+ IV fluids | 3 hours | 67.10% | 6.46% | 5.64% | -0.82% (-1.87%,+0.25%) | -0.25% (-0.57%,+0.08%) |
| Septic shock (N=5,309). 2016-17 mortality: 24.68% |  |  |  |  |  |  |
| Broad-spectrum antibiotics | 1 hour | 23.70% | 25.02% | 23.58% | -1.44% (-4.28%,+1.24%) | -1.09% (-3.26%,+0.95%) |
| Broad-spectrum antibiotics | 3 hours | 74.70% | 30.53% | 22.80% | -7.74% (-10.83%,-4.94%)<sup>d</sup> | -1.88% (-2.61%,-1.19%)<sup>d</sup> |
| IV fluids | 1 hour | 61.80% | 26.91% | 23.34% | -3.58% (-6.01%,-1.17%)<sup>d</sup> | -1.34% (-2.24%,-0.44%)<sup>d</sup> |
| IV fluids | 3 hours | 90.40% | 34.34% | 23.70% | -10.65% (-15.46%,-6.21%)<sup>d</sup> | -0.98% (-1.42%,-0.57%)<sup>d</sup> |
| Broad-spectrum antibiotics<br>+ IV fluids | 1 hour | 21.40% | 25.03% | 23.38% | -1.65% (-4.48%,+1.14%) | -1.29% (-3.53%,+0.90%) |
| Broad-spectrum antibiotics<br>+ IV fluids | 3 hours | 71.80% | 30.15% | 22.63% | -7.52% (-10.54%,-4.93%)<sup>d</sup> | -2.04% (-2.87%,-1.32%)<sup>d</sup> |

CI=confidence interval; ED=emergency department; ED-A=ED arrival; IV=intravenous
<sup>a</sup>2016-2017; <sup>b</sup>Any admission where (1) severe sepsis clinical criteria were met while in the ED, and (2) patient was treated with broad-spectrum antibiotics and IV fluids (any volume) at some point during hospitalization; <sup>c</sup>Any volume; <sup>d</sup>Statistically significant (p<0.05)

**Table 6.**
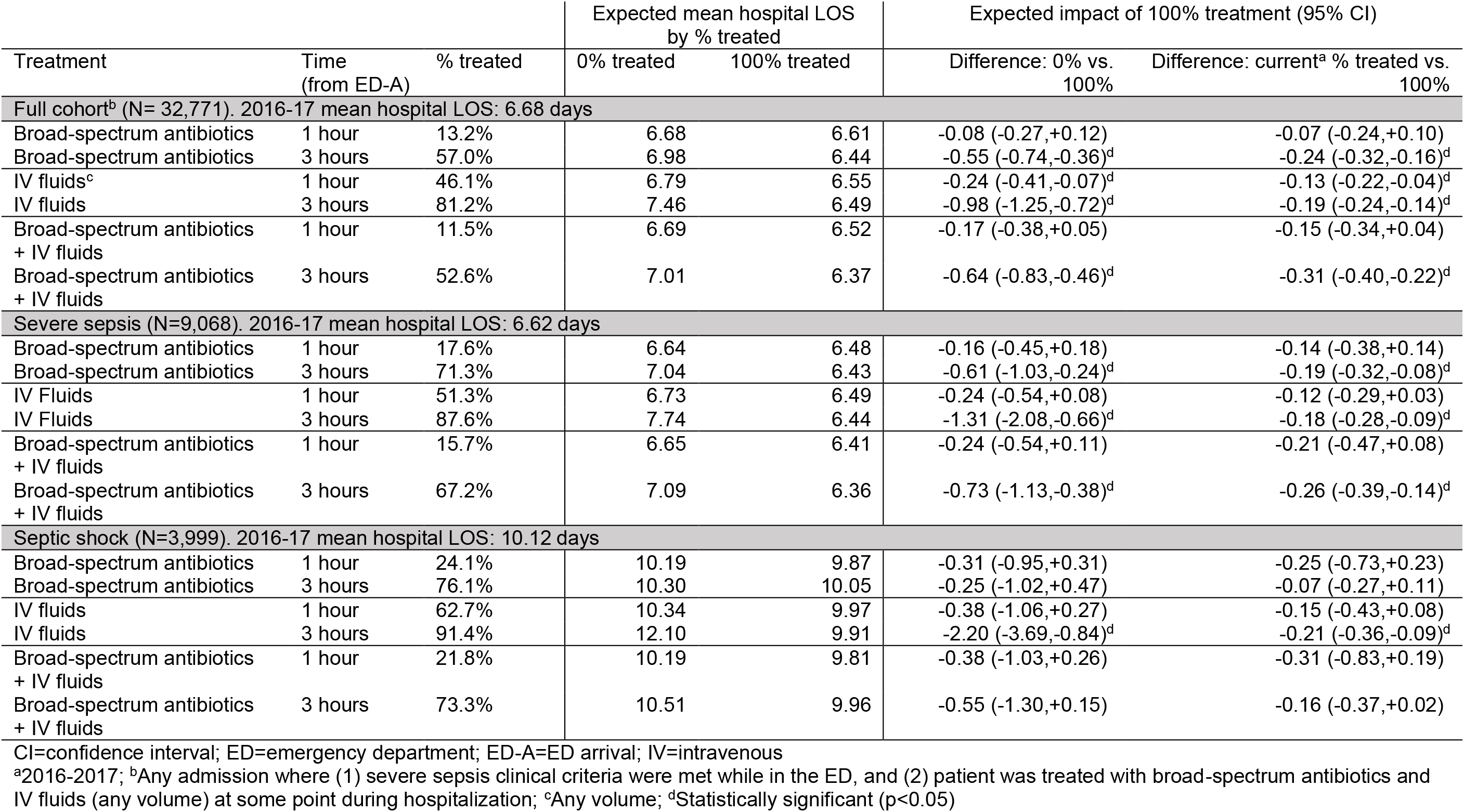
Estimated impact of timely treatment on mean hospital LOS (days) for patients discharged alive, measured from ED arrival.

**Table 7.** Estimated impact of timely treatment on hours in the ICU, measured from ED arrival.

|  |  |  | Expected mean ICU hours<br>by % treated |  | Expected impact of 100% treatment (95% CI) |  |
| --- | --- | --- | --- | --- | --- | --- |
| Treatment | Time<br>(from ED-A) | % treated | 0% treated | 100% treated | Difference: 0% vs. 100% | Difference: current<sup>a</sup> % treated vs.<br>100% |
| Full cohort<sup>b</sup> (N= 10,544). 2016-17 mean time in ICU: 89.43 hours |  |  |  |  |  |  |
| Broad-spectrum antibiotics | 1 hour | 17.6% | 89.65 | 88.25 | -1.41 (-6.85,+4.25) | -1.18 (-5.71,+3.51) |
| Broad-spectrum antibiotics | 3 hours | 61.0% | 93.12 | 86.88 | -6.24 (-11.93,-1.29)<sup>d</sup> | -2.55 (-4.83,-0.58)<sup>d</sup> |
| IV fluids<sup>c</sup> | 1 hour | 54.6% | 93.38 | 86.07 | -7.31 (-12.08,-3.02)<sup>d</sup> | -3.36 (-5.55,-1.44)<sup>d</sup> |
| IV fluids | 3 hours | 84.0% | 109.33 | 85.55 | -23.78 (-32.35,-16.09)<sup>d</sup> | -3.88 (-5.31,-2.64)<sup>d</sup> |
| Broad-spectrum antibiotics<br>+ IV fluids | 1 hour | 15.3% | 89.98 | 86.19 | -3.79 (-9.22,+2.13) | -3.24 (-7.87,+1.79) |
| Broad-spectrum antibiotics<br>+ IV fluids | 3 hours | 56.6% | 94.58 | 85.25 | -9.33 (-14.60,-4.73)<sup>d</sup> | -4.18 (-6.49,-2.10)<sup>d</sup> |
| Severe sepsis (N=2,395). 2016-17 mean time in ICU: 76.24 hours |  |  |  |  |  |  |
| Broad-spectrum antibiotics | 1 hour | 21.9% | 75.70 | 75.73 | +0.03 (-7.68,+9.31) | -0.51 (-6.95,+6.83) |
| Broad-spectrum antibiotics | 3 hours | 72.4% | 83.88 | 72.50 | -11.38 (-22.09,-1.62)<sup>d</sup> | -3.74 (-6.74,-0.79)<sup>d</sup> |
| IV Fluids | 1 hour | 55.2% | 81.78 | 70.50 | -11.28 (-18.13,-3.97)<sup>d</sup> | -5.74 (-9.43,-2.12)<sup>d</sup> |
| IV Fluids | 3 hours | 86.6% | 92.07 | 73.01 | -19.06 (-33.18,-5.34)<sup>d</sup> | -3.23 (-5.54,-1.1)<sup>d</sup> |
| Broad-spectrum antibiotics<br>+ IV fluids | 1 hour | 19.0% | 76.36 | 72.82 | -3.54 (-11.91,+5.64) | -3.42 (-10.02,+4.27) |
| Broad-spectrum antibiotics<br>+ IV fluids | 3 hours | 67.0% | 85.66 | 70.69 | -14.98 (-24.69,-5.44)<sup>d</sup> | -5.55 (-8.9,-1.98)<sup>d</sup> |
| Septic shock (N=3,488). 2016-17 mean time in ICU: 109.93 hours |  |  |  |  |  |  |
| Broad-spectrum antibiotics | 1 hour | 24.8% | 110.13 | 108.61 | -1.51 (-11.37,+9.36) | -1.32 (-8.9,+6.77) |
| Broad-spectrum antibiotics | 3 hours | 76.5% | 110.22 | 109.61 | -0.61 (-12.08,+10.43) | -0.32 (-3.4,+2.39) |
| IV fluids | 1 hour | 63.1% | 111.12 | 108.93 | -2.20 (-12.92,+6.81) | -1.01 (-5.05,+2.27) |
| IV fluids | 3 hours | 91.5% | 142.24 | 106.57 | -35.66 (-60.29,-14.5)<sup>d</sup> | -3.36 (-5.95,-1.52)<sup>d</sup> |
| Broad-spectrum antibiotics<br>+ IV fluids | 1 hour | 22.4% | 110.68 | 106.49 | -4.19 (-13.93,+6.15) | -3.45 (-11.06,+4.36) |
| Broad-spectrum antibiotics<br>+ IV fluids | 3 hours | 73.7% | 115.29 | 107.63 | -7.66 (-18.19,+3.38) | -2.3 (-5.34,+0.74) |

CI=confidence interval; ED=emergency department; ED-A=ED arrival; IV=intravenous
Analysis includes patients discharged alive who went to the ICU at some point during hospitalization.
<sup>a</sup>2016-2017; <sup>b</sup>Any admission where (1) severe sepsis clinical criteria were met while in the ED, and (2) patient was treated with broad-spectrum antibiotics and IV fluids (any volume) at some point during hospitalization; <sup>c</sup>Any volume; <sup>d</sup>Statistically significant (p<0.05)

**Table 8.**
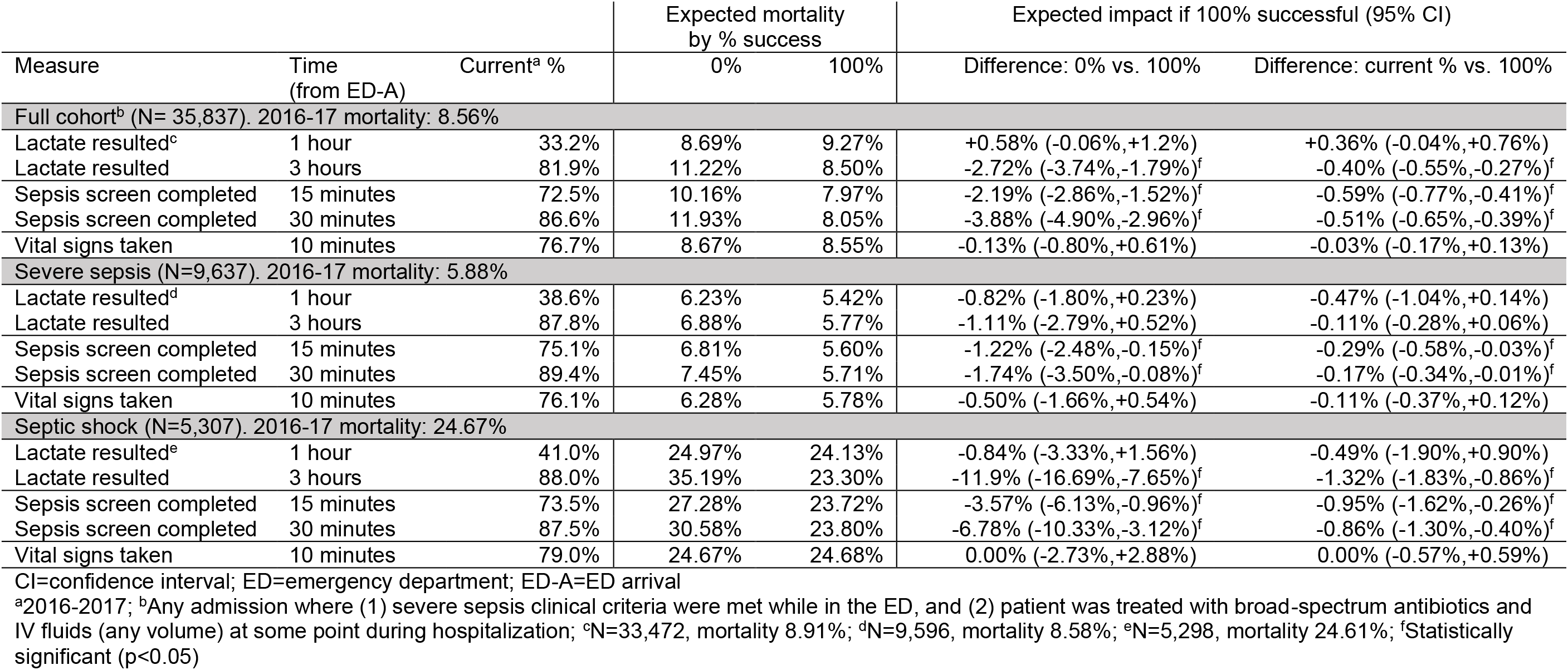
Estimated impact of timely lactate measurement, sepsis screening, and vital signs on in-hospital mortality.

**Table 9.** Estimated impact of timely lactate measurement, sepsis screening, and vital signs on hospital length of stay (days) for patients discharged alive.

|  |  |  | Expected mean hospital LOS<br>by % success |  | Expected impact if 100% successful (95% CI) |  |
| --- | --- | --- | --- | --- | --- | --- |
| Measure | Time<br>(from ED-A) | Current<sup>a</sup> % | 0% | 100% | Difference: 0% vs. 100% | Difference: current % vs. 100% |
| Full cohort<sup>b</sup> (N= 32,767). 2016-17 mean hospital LOS: 6.68 days |  |  |  |  |  |  |
| Lactate resulted<sup>c</sup> | 1 hour | 32.6% | 6.78 | 6.69 | -0.10 (-0.28,+0.09) | -0.07 (-0.19,+0.06) |
| Lactate resulted | 3 hours | 82.0% | 7.70 | 6.54 | -1.17 (-1.48,-0.87)<sup>f</sup> | -0.22 (-0.27,-0.16)<sup>f</sup> |
| Sepsis screen completed | 15 minutes | 72.9% | 6.77 | 6.64 | -0.13 (-0.32,+0.04) | -0.04 (-0.09,+0.01) |
| Sepsis screen completed | 30 minutes | 87.1% | 7.09 | 6.61 | -0.48 (-0.78,-0.21)<sup>f</sup> | -0.07 (-0.11,-0.03)<sup>f</sup> |
| Vital signs taken | 10 minutes | 76.5% | 6.57 | 6.71 | 0.14 (-0.05,+0.33) | 0.03 (-0.01,+0.08) |
| Severe sepsis (N=9,068). 2016-17 mean hospital LOS: 6.62 days |  |  |  |  |  |  |
| Lactate resulted<sup>d</sup> | 1 hour | 38.6% | 6.71 | 6.46 | -0.25 (-0.54,+0.07) | -0.16 (-0.34,+0.04) |
| Lactate resulted | 3 hours | 87.8% | 7.45 | 6.49 | -0.96 (-1.61,-0.28)<sup>f</sup> | -0.13 (-0.22,-0.04)<sup>f</sup> |
| Sepsis screen completed | 15 minutes | 75.2% | 6.59 | 6.62 | +0.02 (-0.31,+0.33) | 0.00 (-0.08,+0.07) |
| Sepsis screen completed | 30 minutes | 89.5% | 6.82 | 6.59 | -0.23 (-0.70,+0.22) | -0.03 (-0.08,+0.01) |
| Vital signs taken | 10 minutes | 76.0% | 6.42 | 6.67 | +0.26 (-0.08,+0.54) | 0.06 (-0.03,+0.12) |
| Septic shock (N=3,998). 2016-17 mean hospital LOS: 10.12 days |  |  |  |  |  |  |
| Lactate resulted<sup>e</sup> | 1 hour | 40.7% | 10.12 | 10.13 | +0.01 (-0.59,+0.61) | 0.00 (-0.37,+0.35) |
| Lactate resulted | 3 hours | 89.1% | 11.2 | 9.97 | -1.22 (-2.45,-0.13)<sup>f</sup> | -0.15 (-0.30,-0.03)<sup>f</sup> |
| Sepsis screen completed | 15 minutes | 74.6% | 9.85 | 10.21 | +0.36 (-0.31,+1.02) | +0.09 (-0.10,+0.25) |
| Sepsis screen completed | 30 minutes | 88.6% | 10.02 | 10.13 | +0.11 (-0.69,+0.91) | +0.01 (-0.11,+0.10) |
| Vital signs taken | 10 minutes | 78.9% | 9.68 | 10.24 | +0.56 (-0.12,+1.21) | +0.12 (-0.04,+0.26) |

CI=confidence interval; ED=emergency department; ED-A=ED arrival; IV=intravenous
<sup>a</sup>2016-2017; <sup>b</sup>Any admission where (1) severe sepsis clinical criteria were met while in the ED, and (2) patient was treated with broad-spectrum antibiotics and IV fluids (any volume) at some point during hospitalization; <sup>c</sup>N=30,489, mean LOS 6.75 days; <sup>d</sup>N=9,029, mean LOS 6.62 days; <sup>e</sup>N=3,994, mean LOS 10.13 days; <sup>f</sup>Statistically significant (p<0.05)

**Table 10.** Estimated impact of timely lactate measurement and sepsis screening on hours spent in the ICU.

|  |  |  | Expected mean ICU hours<br>by % success |  | Expected impact if 100% successful (95% CI) |  |
| --- | --- | --- | --- | --- | --- | --- |
| Measure | Time<br>(from ED-A) | Current<sup>a</sup> % | 0% | 100% | Difference: 0% vs. 100% | Difference: current % vs. 100% |
| Full cohort<sup>b</sup> (N=10,543). 2016-17 mean time in ICU: 89.44 hours |  |  |  |  |  |  |
| Lactate resulted<sup>c</sup> | 1 hour | 37.1% | 88.45 | 93.24 | +4.79 (+0.13,+9.69)<sup>f</sup> | +3.06 (+0.09,+6.17)<sup>f</sup> |
| Lactate resulted | 3 hours | 83.2% | 106.88 | 86.73 | -20.15 (-27.95,-11.91)<sup>f</sup> | -3.46 (-4.90,-2.09)<sup>f</sup> |
| Sepsis screen completed | 15 minutes | 71.7% | 93.69 | 87.64 | -6.04 (-10.83,-1.46)<sup>f</sup> | -1.80 (-3.21,-0.47)<sup>f</sup> |
| Sepsis screen completed | 30 minutes | 84.9% | 95.22 | 88.29 | -6.93 (-13.18,-1.60)<sup>f</sup> | -1.15 (-2.20,-0.28)<sup>f</sup> |
| Vital signs taken | 10 minutes | 79.4% | 89.01 | 89.52 | +0.51 (-5.13,+5.87) | +0.09 (-1.11,+1.18) |
| Severe sepsis (N=2,395). 2016-17 mean time in ICU: 76.24 hours |  |  |  |  |  |  |
| Lactate resulted<sup>d</sup> | 1 hour | 39.2% | 72.28 | 81.34 | +9.06 (+1.38,+16.49)<sup>f</sup> | +5.16 (+0.25,+9.97)<sup>f</sup> |
| Lactate resulted | 3 hours | 87.6% | 93.18 | 73.2 | -19.99 (-35.00,-3.97)<sup>f</sup> | -2.99 (-5.74,-0.69)<sup>f</sup> |
| Sepsis screen completed | 15 minutes | 74.1% | 78.81 | 74.61 | -4.2 (-12.91,+3.42) | -1.63 (-4.10,+0.34) |
| Sepsis screen completed | 30 minutes | 87.5% | 81.3 | 74.89 | -6.41 (-18.3,+2.49) | -1.35 (-3.25,-0.03)<sup>f</sup> |
| Vital signs taken | 10 minutes | 78.8% | 73.54 | 76.26 | +2.72 (-6.32,+10.66) | 0.02 (-2.08,+1.73) |
| Septic shock (N=3,487). 2016-17 mean time in ICU: 109.96 hours |  |  |  |  |  |  |
| Lactate resulted<sup>e</sup> | 1 hour | 40.4% | 104.28 | 118.35 | +14.07 (+5.32,+23.41)<sup>f</sup> | +8.39 (+2.96,+13.99)<sup>f</sup> |
| Lactate resulted | 3 hours | 89.3% | 121.61 | 108.27 | -13.34 (-32.92,+4.97) | -1.69 (-4.15,+0.29) |
| Sepsis screen completed | 15 minutes | 74.5% | 111.11 | 109.33 | -1.78 (-13.21,+8.84) | -0.64 (-3.71,+2.00) |
| Sepsis screen completed | 30 minutes | 88.5% | 107.24 | 110.15 | +2.91 (-10.32,+14.45) | +0.18 (-1.63,+1.57) |
| Vital signs taken | 10 minutes | 79.1% | 111.32 | 109.35 | -1.97 (-13.22,+9.31) | -0.59 (-3.22,+1.86) |

CI=confidence interval; ED=emergency department; ED-A=ED arrival; IV=intravenous
Analysis includes patients discharged alive who went to the ICU at some point during hospitalization.
<sup>a</sup>2016-2017; <sup>b</sup>Any admission where (1) severe sepsis clinical criteria were met while in the ED, and (2) patient was treated with broad-spectrum antibiotics and IV fluids (any volume) at some point during hospitalization; <sup>c</sup>N=10,113, mean ICU hours 90.19; <sup>d</sup>N=2,390, mean ICU hours 76.18; <sup>e</sup>N=3,484, mean ICU hours 109.96; <sup>f</sup>Statistically significant (p<0.05)

### Mortality

All versions of treatment within 1 and 3 hours of sepsis onset (Table 2) were found to have a positive impact on mortality, with estimated reductions in mortality rate ranging from 1.01% (antibiotics within 1 hour) to 2.85% (fluids within 1 hour) for the full cohort (N=35,847, mortality rate 8.58%), 1.03% (antibiotics within 1 hour) and 2.94% (fluids within 1 hour) for severe sepsis (N=9,638, mortality rate 5.89%), and from 5.80% (antibiotics within 1 hour) and 14.66% (fluids within 3 hours) for septic shock (N=5,309, mortality rate 24.68%). For all three cohorts, timely initiation of IV fluids was associated with the largest estimated mortality rate reduction.

The greatest opportunity for improvement in current practice, however, was not in the area of IV fluid initiation, because the vast majority of patients (89.8%) were already receiving fluids within 3 hours of sepsis onset (or prior to sepsis onset). The biggest potential gains were estimated to be in the area of increasing the combined initiation of antibiotics and fluids within 1 hour of sepsis onset – only 46.6% of patients (57.3% severe sepsis, 56.7% septic shock) received both treatments within this time frame. For all three cohorts, this combined treatment within 1 hour was associated with the largest mortality reduction as compared to current practice (0.8% for the full cohort, 0.77% for severe sepsis, and 2.94% for septic shock).

Looking at treatment within 1 and 3 hours of ED arrival (Table 5), estimated mortality rate reductions were lower, and less likely to be statistically significant (p<0.05). The largest estimated reductions were still in the area of IV fluid initiation, specifically within 3 hours of ED arrival (2.46% full cohort, 1.96% severe sepsis, 10.65% septic shock). The largest estimated impact of changing current practice was in administering antibiotics and fluids within 3 hours of ED arrival (0.49% mortality reduction overall, 2.04% reduction for septic shock).

Table 8 shows the estimated impact of timely lactate measurement, sepsis screening, and vital signs on mortality. Results suggest that taking vital signs within 10 minutes of ED arrival does not impact the mortality rate for any cohort, nor does receiving lactate test results within 1 hour of ED arrival (for those with a lactate result, N=33,742) Lactate results within 3 hours of arrival was associated with a reduction in mortality rate for the full cohort and for septic shock admissions (2.72% and 11.9%, respectively). For the latter, this is the largest estimated impact shown in Table 8. Sepsis screen completion within 30 minutes had the largest estimated impact for the full cohort and for severe sepsis admissions (mortality rate reduction 3.88% and 1.74%, respectively). Despite the high success of early sepsis screening (72.5% within 15 minutes, 86.6% within 30 minutes) and of lactate measurement within 3 hours (81.9%), these areas still showed the largest estimated impacts of a change to current practice, particularly within those who developed septic shock. Within this population, mortality rates could be reduced by an estimated 1.32% by returning lactate results within 3 hours in all cases, and by 0.95% if all sepsis screens were documented within 15 minutes of ED arrival. Within the full cohort, sepsis screening within 15 minutes was also the greatest identified opportunity, with an estimated mortality rate reduction of 0.59% with a change to current practice.

Results displayed in Tables 2, 5, and 8 can also be viewed in the forest plots shown in Figures 2-7.

**Figure 2.**
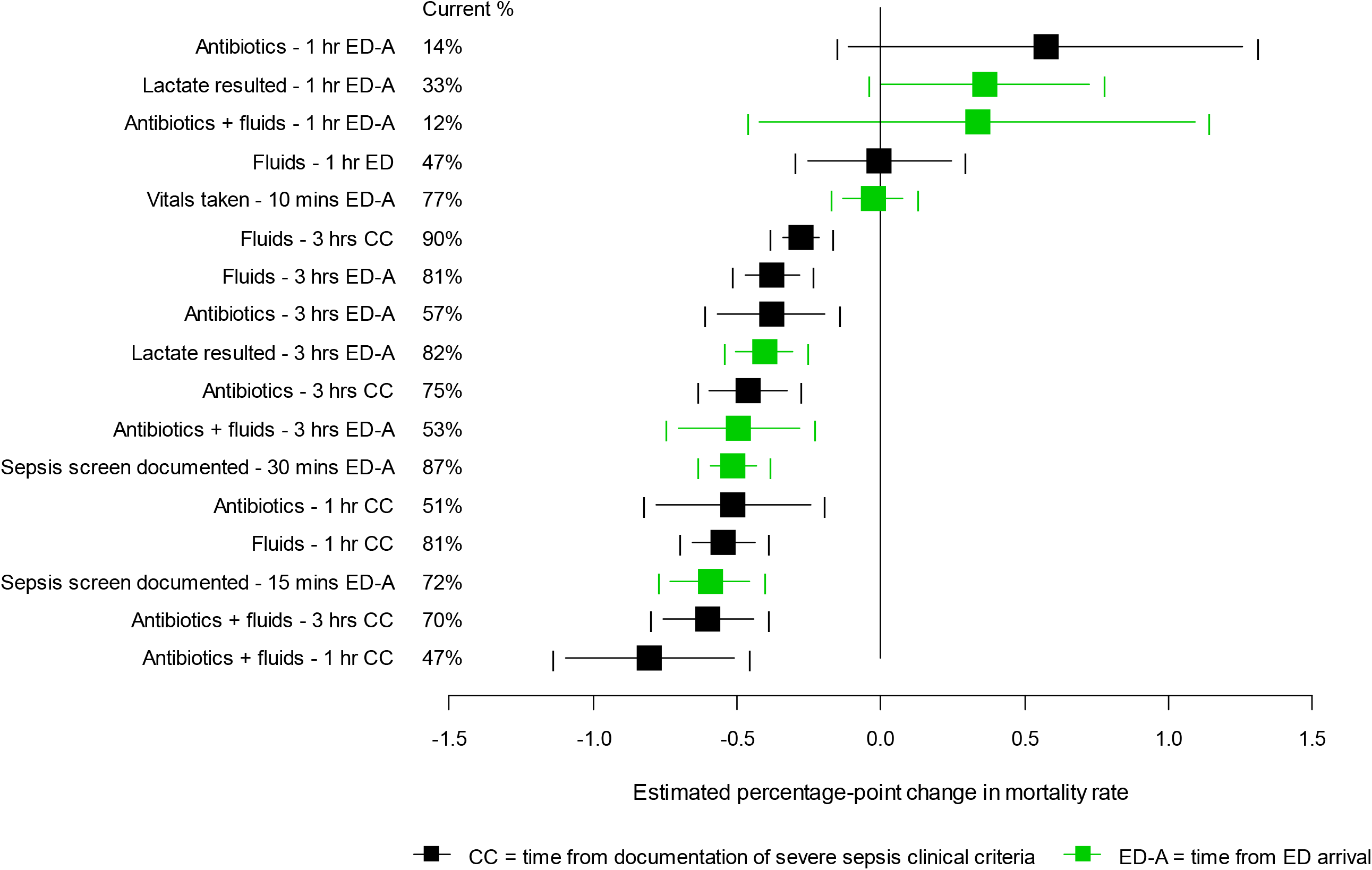
Estimated opportunity to impact in-hospital mortality (100% success vs. current %). Point estimates and 95% confidence intervals are shown. The vertical line at zero indicates no impact. Full cohort; sepsis screen/vital signs N=35,837, lactate resulted N=33,472, otherwise N=35,847. Data detail in Tables 2, 5, and 8.

**Figure 3.**
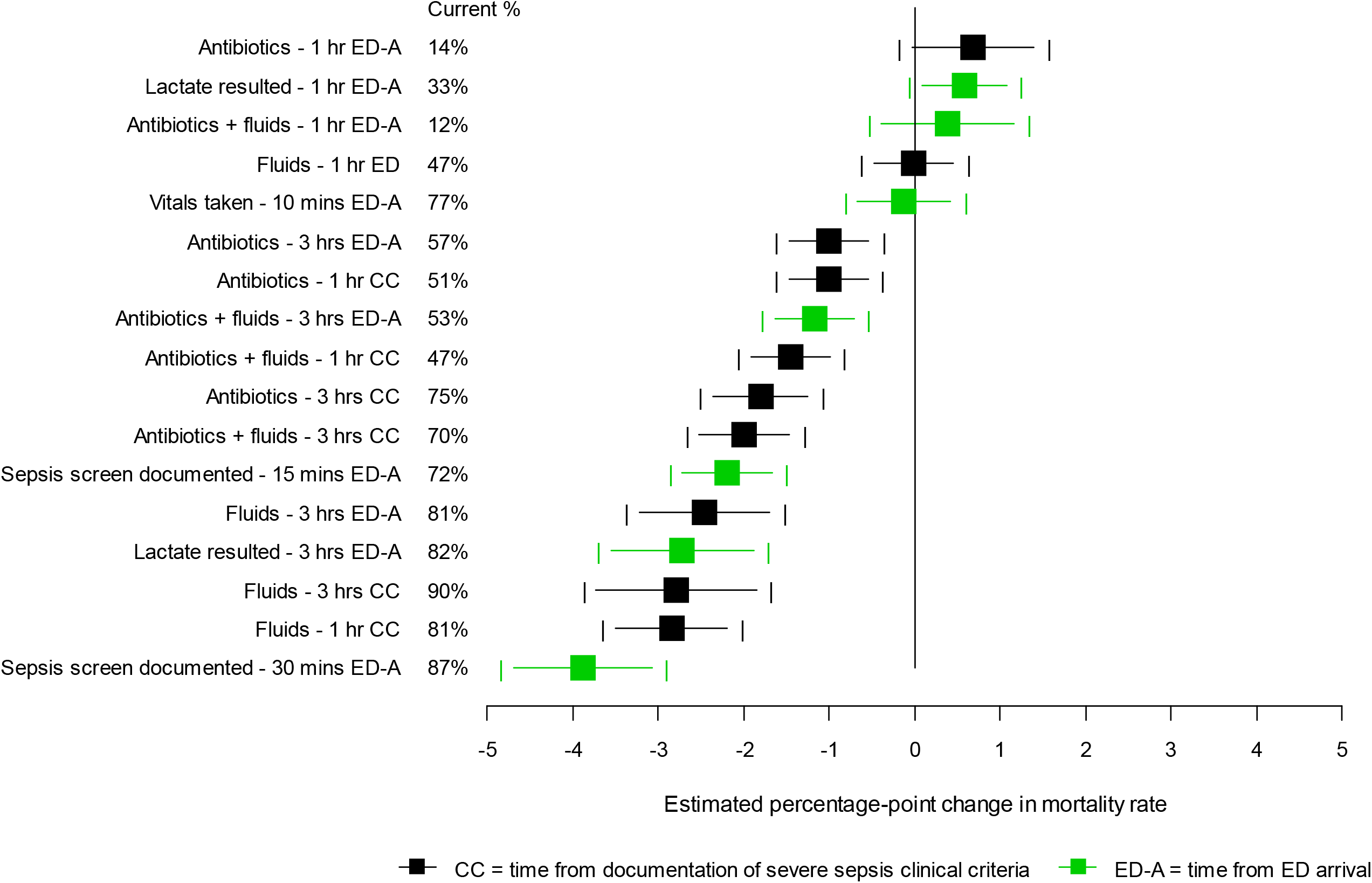
Estimated impact of timely identification and treatment on in-hospital mortality (100% success vs. 0% success). Point estimates and 95% confidence intervals are shown. The vertical line at zero indicates no impact. Full cohort; sepsis screen/vital signs N=35,837, lactate resulted N=33,472, otherwise N=35,847. Data detail in Tables 2, 5, and 8.

**Figure 4.**
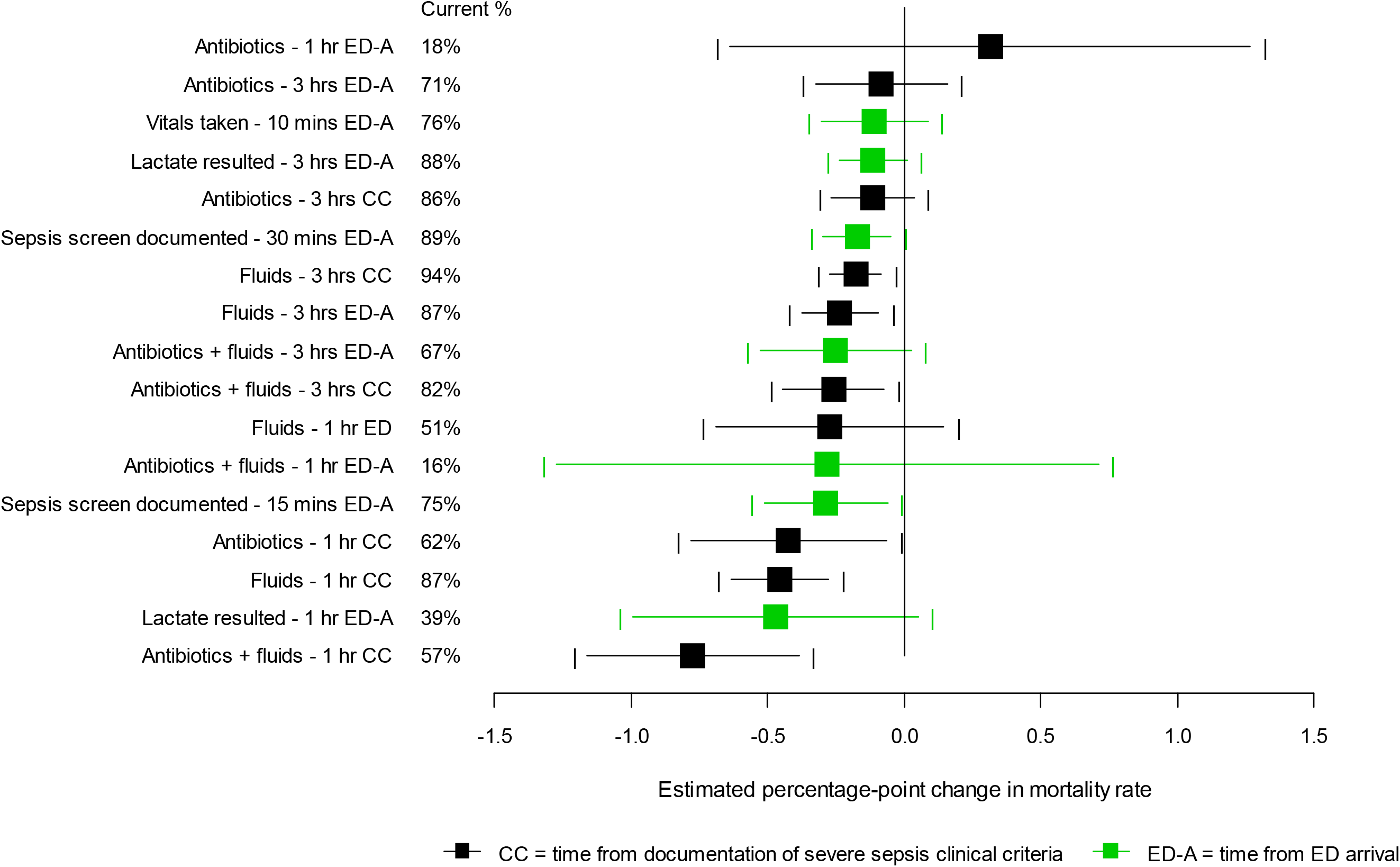
Estimated opportunity to impact in-hospital mortality (100% success vs. current %). Point estimates and 95% confidence intervals are shown. The vertical line at zero indicates no impact. Severe sepsis cohort; sepsis screen/vital signs N=9.637, lactate resulted N=9.596, otherwise N=9.638. Data detail in Tables 2, 5, and 8.

**Figure 5.**
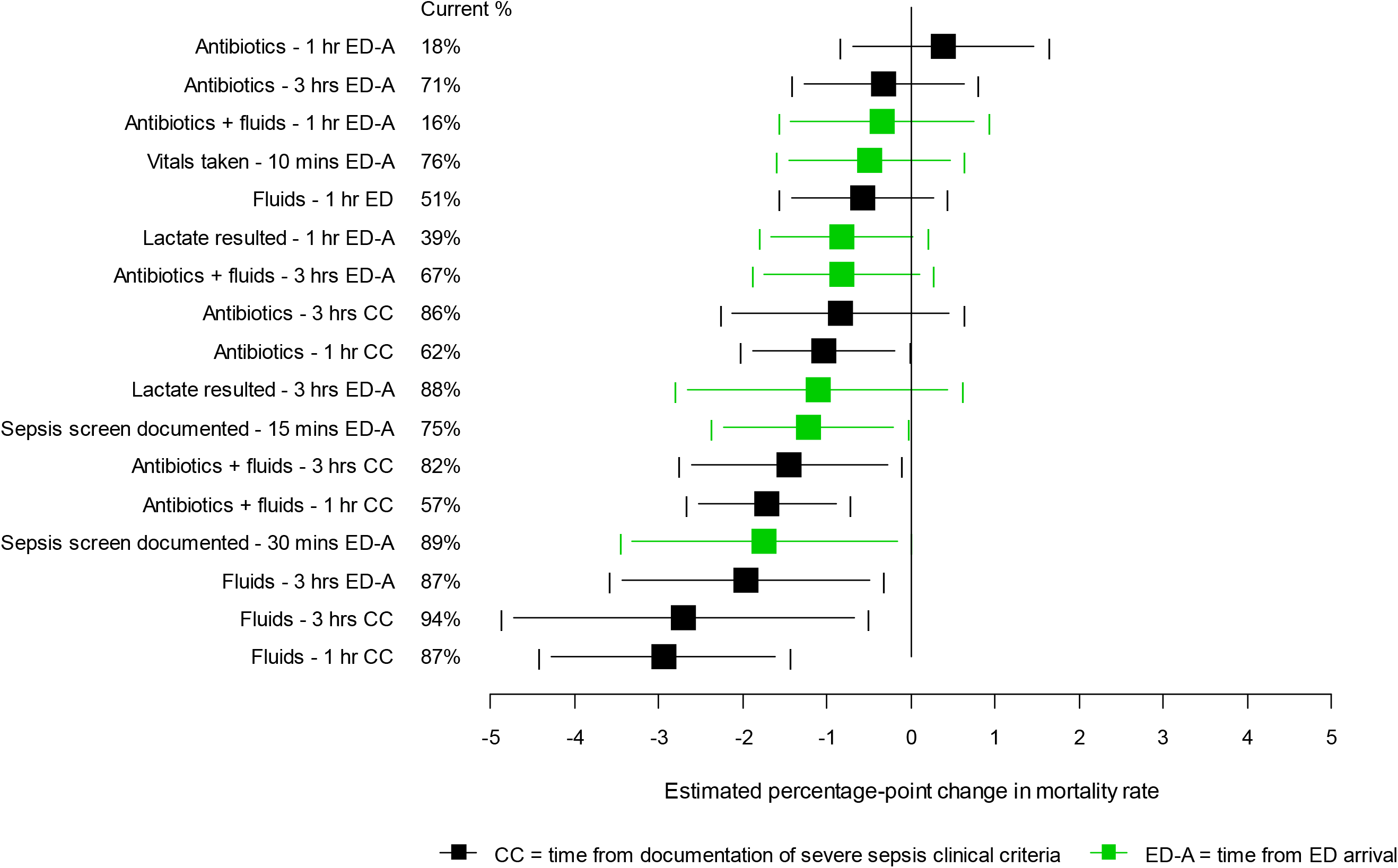
Estimated impact of timely identification and treatment on in-hospital mortality (100% success vs. 0% success). Point estimates and 95% confidence intervals are shown. The vertical line at zero indicates no impact. Severe sepsis cohort; sepsis screen/vital signs N=9.637, lactate resulted N=9.596, otherwise N=9.638. Data detail in Tables 2, 5, and 8.

**Figure 6.**
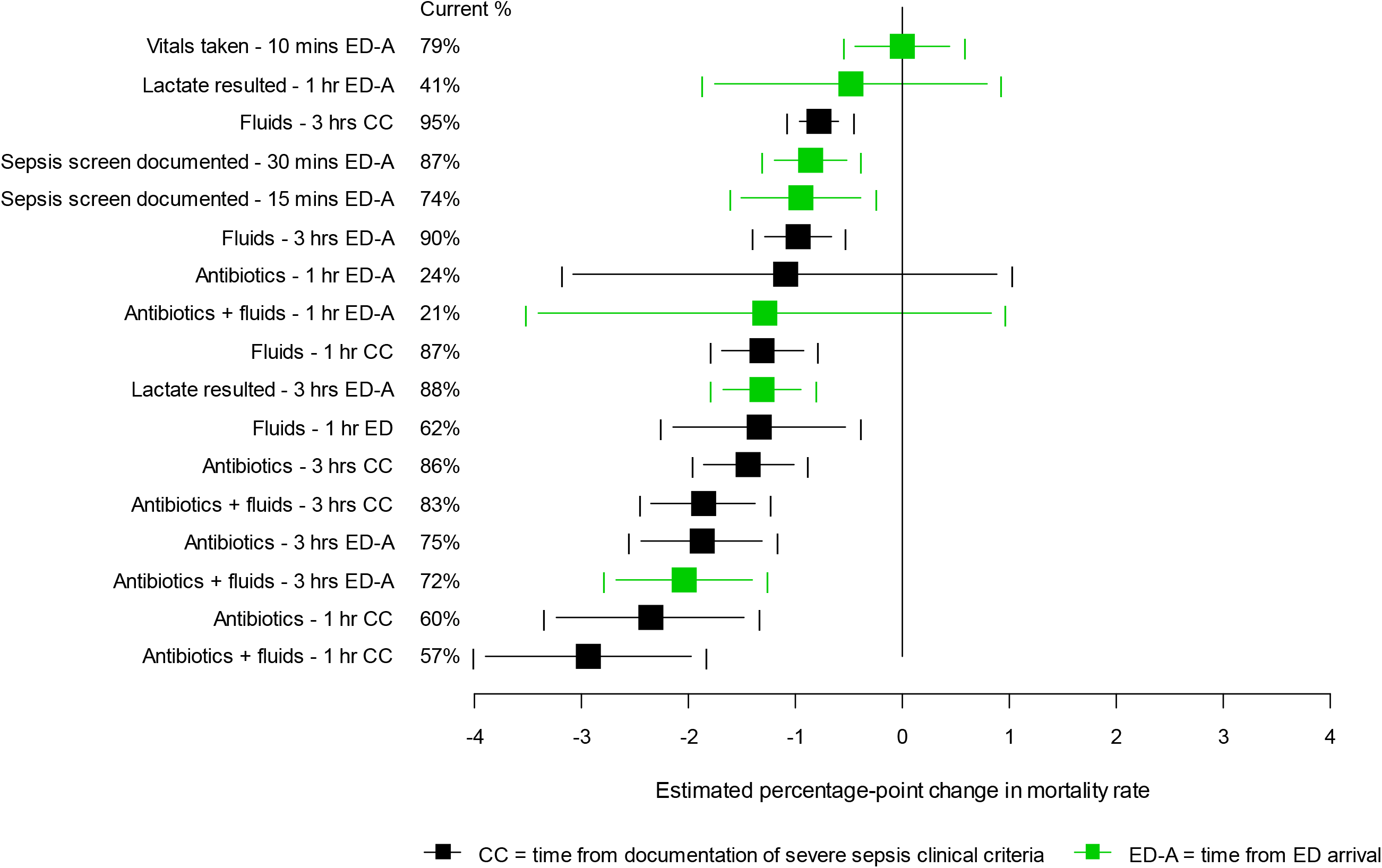
Estimated opportunity to impact in-hospital mortality (100% success vs. current %). Point estimates and 95% confidence intervals are shown. The vertical line at zero indicates no impact. Septic shock cohort; sepsis screen/vital signs N=5,307, lactate resulted N=5,298, otherwise N=5,309. Data detail in Tables 2, 5, and 8.

**Figure 7.**
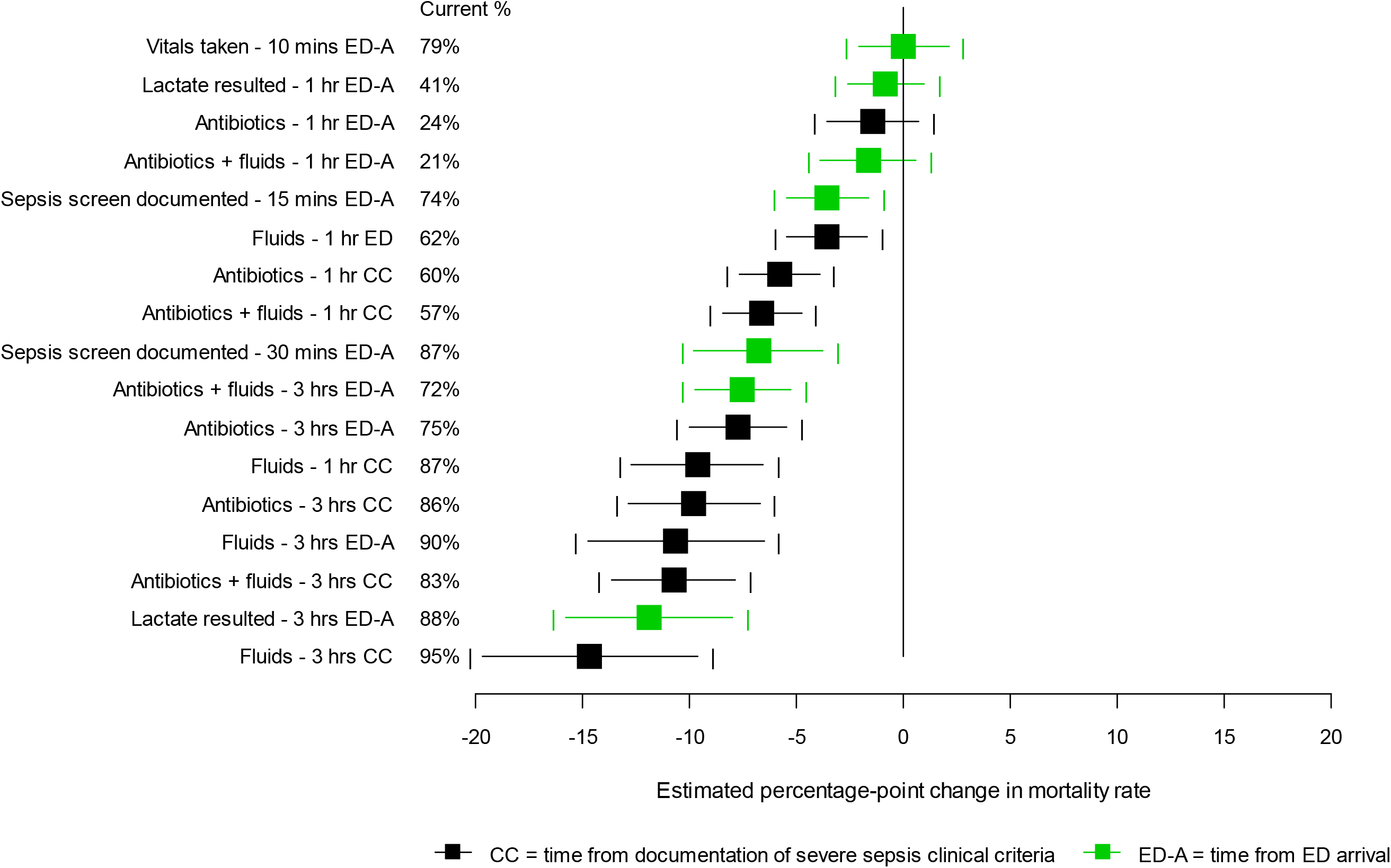
Estimated impact of timely identification and treatment on in-hospital mortality (100% success vs. 0% success). Septic shock cohort; sepsis screen/vital signs N=5,307, lactate resulted N=5,298, otherwise N=5,309. Data detail in Tables 2, 5, and 8.

### Hospital length of stay

For patients discharged alive (N=32,771 full cohort, N=9,068 severe sepsis, N=3,999 septic shock), timely treatment in relation to sepsis onset (Table 3) was associated with statistically significant (p<0.05) reductions in hospital LOS (days). For all three cohorts, the largest estimated impact was associated with IV fluid initiation within 3 hours of sepsis onset – an estimated 1.39 fewer days overall, 2.3 days for severe sepsis admissions, and 3.07 days for septic shock. The next largest reduction in hospital LOS was associated with the combined treatment with antibiotics and IV fluids within 3 hours (1.12 days full cohort, 1.36 days severe sepsis, 1.67 days septic shock). The largest impact associated with a change in current practice was in administering both broad-spectrum antibiotics and IV fluids within 1 or 3 hours, for a modest 0.37- or 0.35-day reduction in overall average hospital LOS.

When treatment time was measured from ED arrival (Table 6), IV fluid initiation within 3 hours produced the largest estimated reductions in LOS (0.98 days full cohort, 1.31 days severe sepsis, 2.20 days septic shock); for the septic shock cohort, this was the only estimated impact in Table 6 whose 95% CI did not cross zero. The largest estimate associated with a change in current practice was for combined initiation of IV fluids and antibiotics within 3 hours of ED arrival (in 2016-17 52.6% of the full cohort received both treatments within this time frame), producing an estimated 0.31-day reduction in average LOS.

Table 9 shows that lactate measurement within 3 hours of ED arrival is associated with reduced hospital LOS, among those with a lactate result (N=30,489; N=9,029 severe sepsis, N=3,994 septic shock). The estimated reductions were 1.17 days for the full cohort, 0.96 days for severe sepsis, and 1.22 days for septic shock. Neither measurement of vital signs within 10 minutes or sepsis screening within 15/30 minutes appeared to impact LOS for severe sepsis and septic shock admissions; for the full cohort, sepsis screening within 30 minutes was associated with a modest (0.48-day) LOS reduction. The results in Table 9 do not reveal opportunities for impactful change to current practice – the largest projected LOS reduction is in lactate measurement within 3 hours for the full cohort, associated with an estimated 0.22-day lower average LOS. The results in tables 3, 6, and 9 are compared graphically for the three cohorts in Figures 8-13.

**Figure 8.**
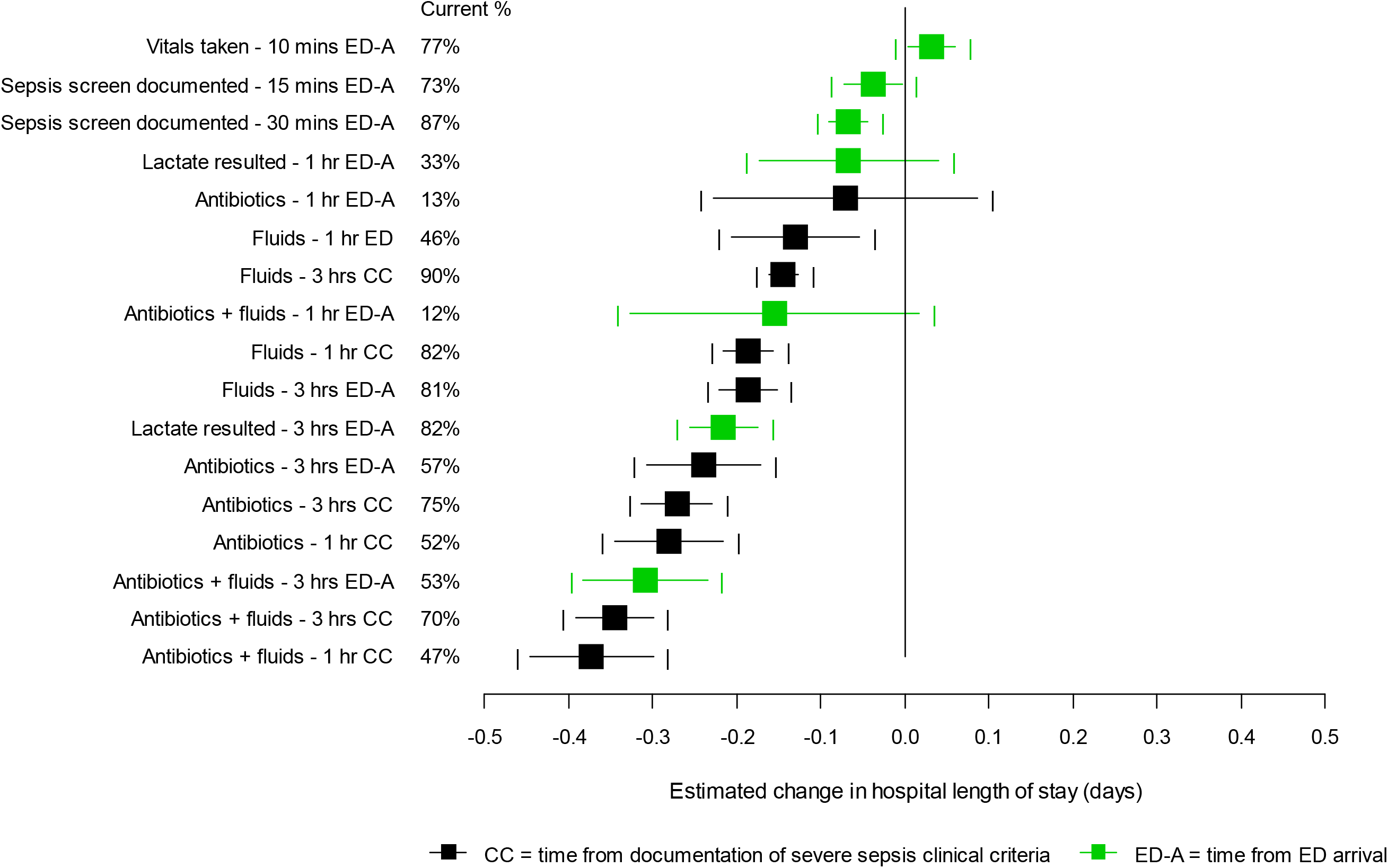
Estimated opportunity to impact hospital length of stay (days) for patients discharged alive (100% success vs. current %). Point estimates and 95% confidence intervals are shown. The vertical line at zero indicates no impact. Full cohort; sepsis screen/vital signs N=32,767, lactate resulted N=30,489, otherwise N=32,771. Data detail in Tables 3, 6, and 9.

**Figure 9.**
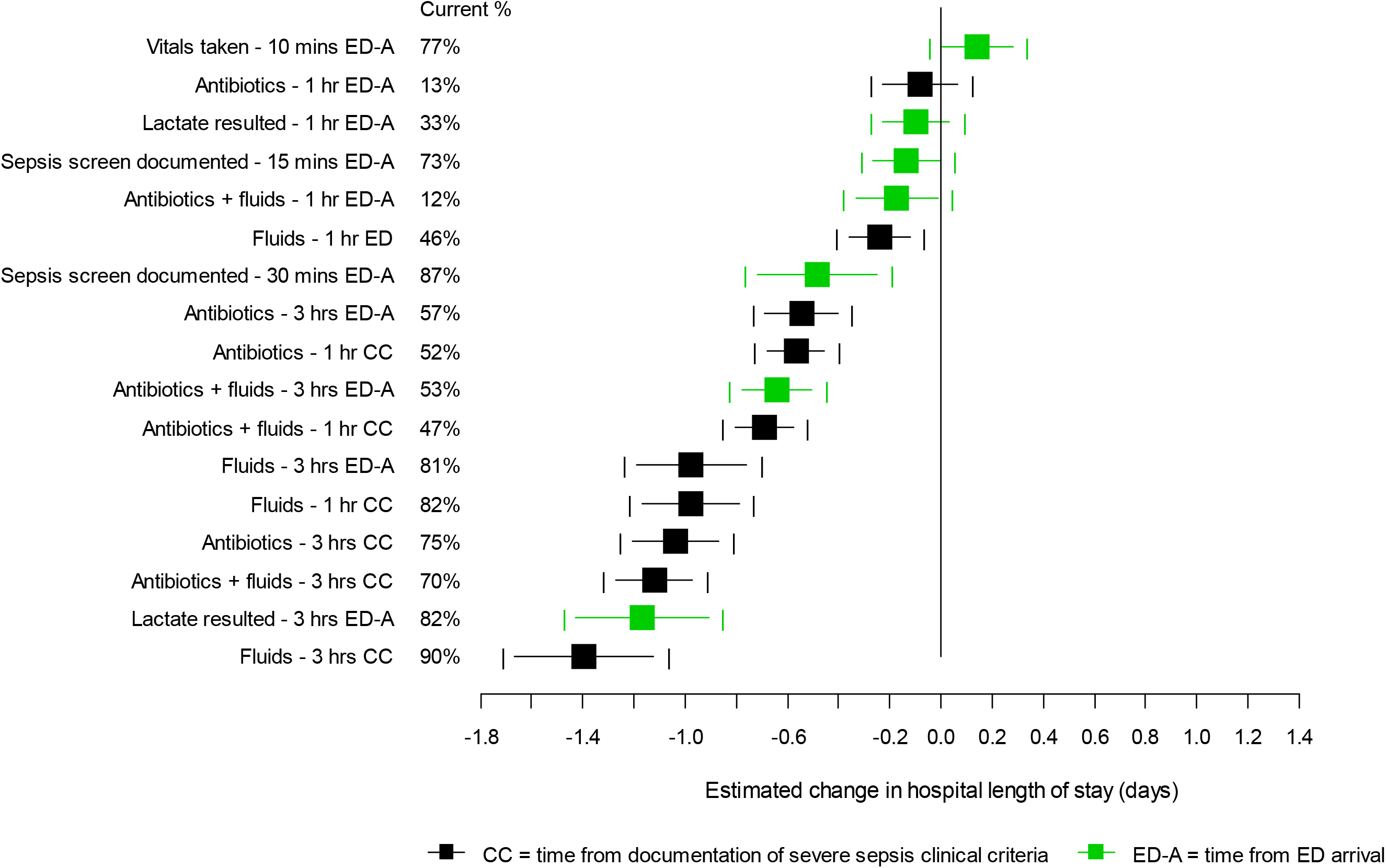
Estimated impact of timely identification and treatment on hospital length of stay (days) for patients discharged alive (100% success vs. 0% success). Point estimates and 95% confidence intervals are shown. The vertical line at zero indicates no impact. Full cohort; sepsis screen/vital signs N=32,767, lactate resulted N=30,489, otherwise N=32,771. Data detail in Tables 3, 6, and 9.

**Figure 10.**
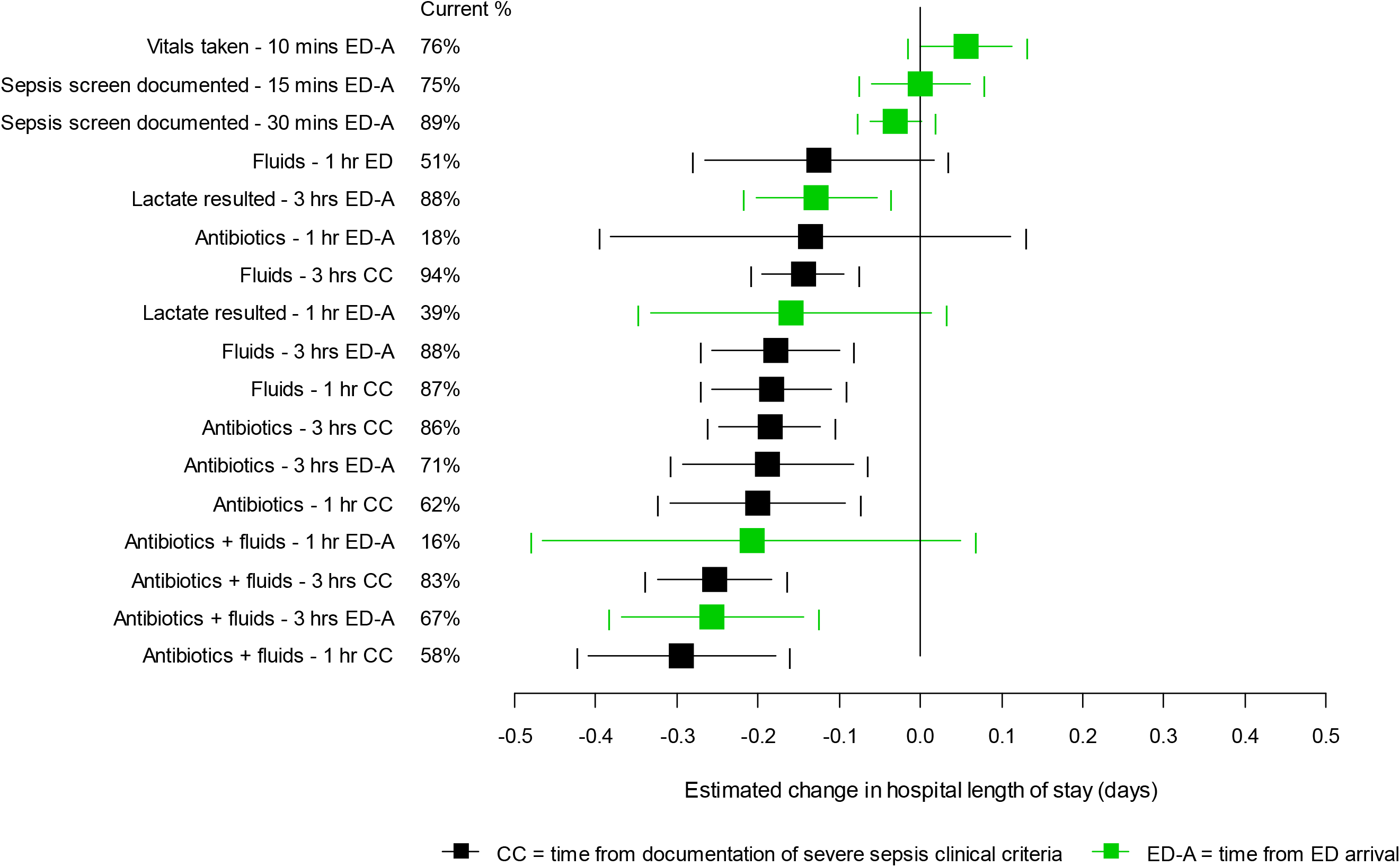
Estimated opportunity to impact hospital length of stay (days) for patients discharged alive (100% success vs. current %). Point estimates and 95% confidence intervals are shown. The vertical line at zero indicates no impact. Severe sepsis cohort; lactate resulted N=9,029, otherwise N=9.068. Data detail in Tables 3, 6, and 9.

**Figure 11.**
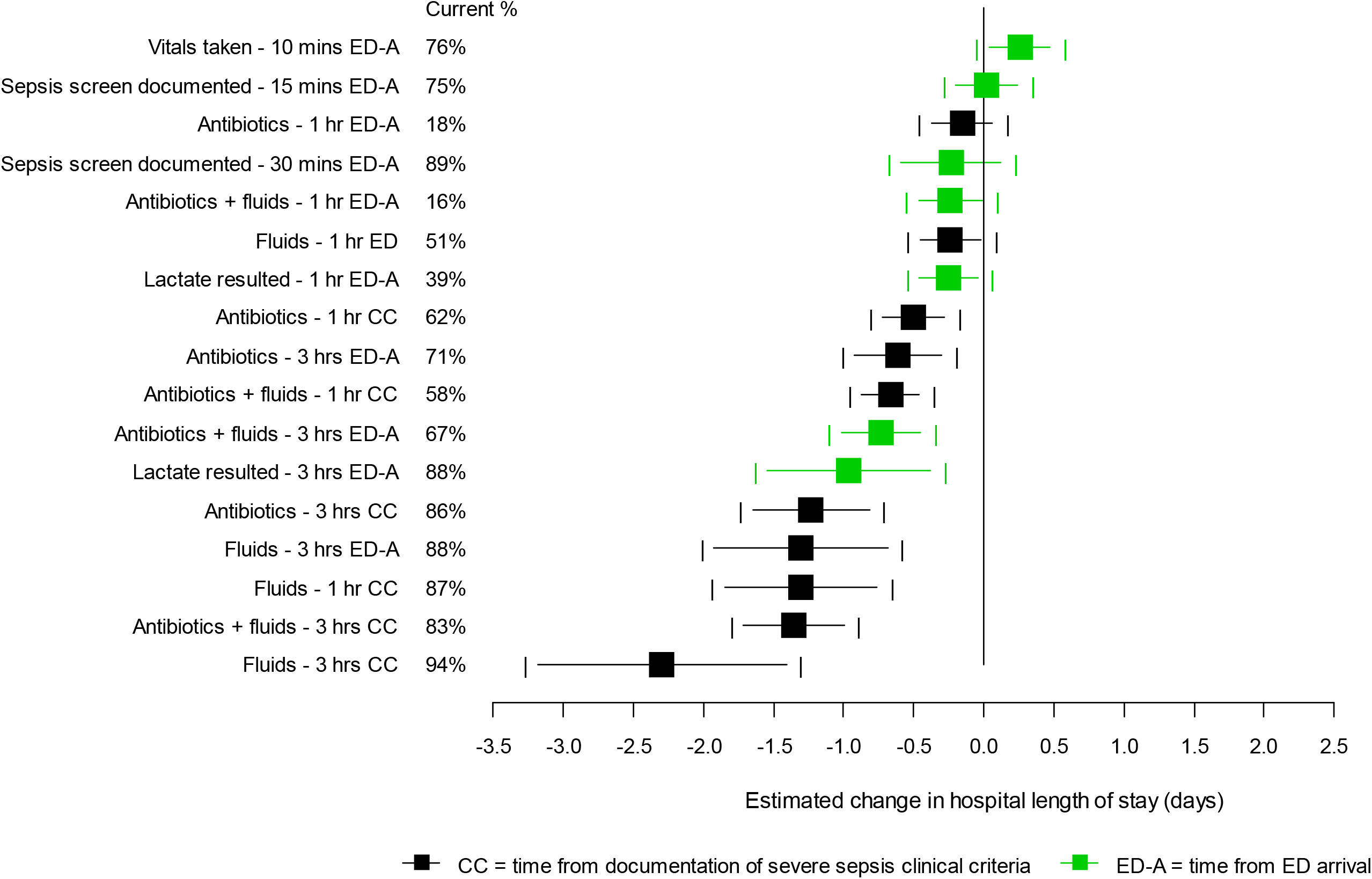
Estimated impact of timely identification and treatment on hospital length of stay (days) for patients discharged alive (100% success vs. 0% success). Point estimates and 95% confidence intervals are shown. The vertical line at zero indicates no impact. Severe sepsis cohort; lactate resulted N=9,029, otherwise N=9.068. Data detail in Tables 3, 6, and 9.

**Figure 12.**
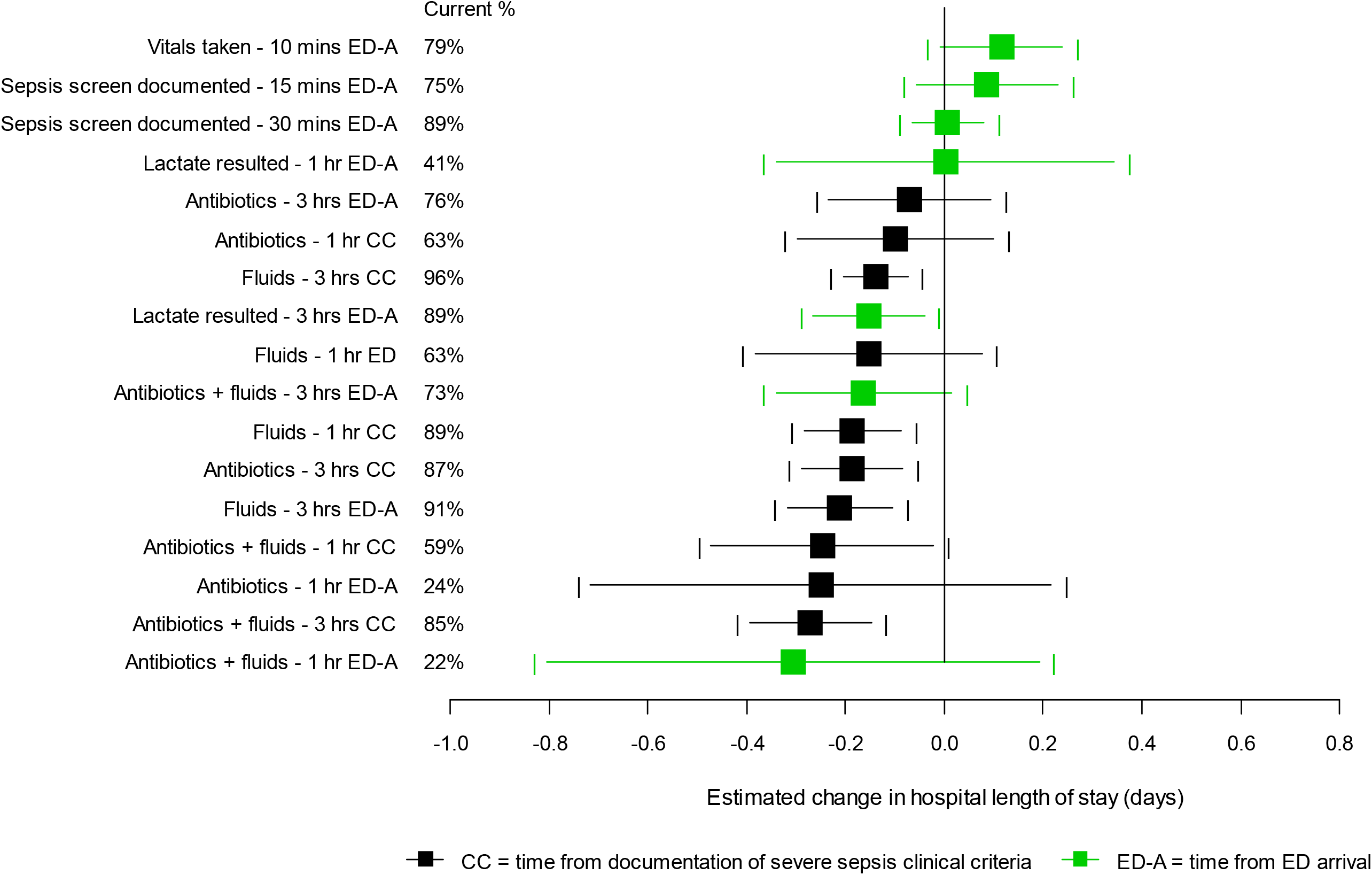
Estimated opportunity to impact hospital length of stay (days) for patients discharged alive (100% success vs. current %). Point estimates and 95% confidence intervals are shown. The vertical line at zero indicates no impact. Septic shock cohort; sepsis screen/vital signs N=3,998, lactate resulted N=3,994, otherwise N=3,999. Data detail in Tables 3, 6, and 9.

**Figure 13.**
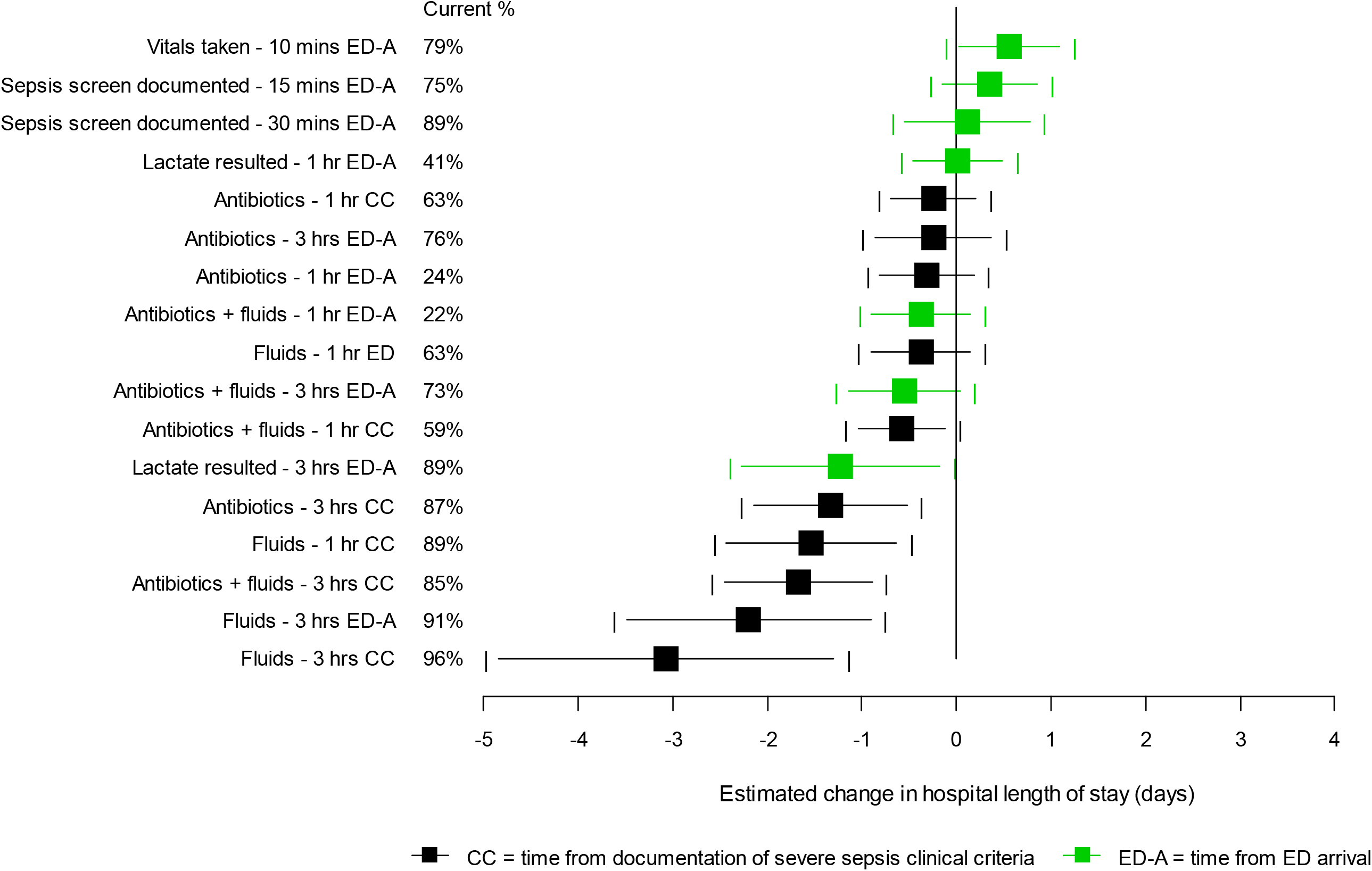
Estimated impact of timely identification and treatment on hospital length of stay (days) for patients discharged alive (100% success vs. 0% success). Point estimates and 95% confidence intervals are shown. The vertical line at zero indicates no impact. Septic shock cohort; sepsis screen/vital signs N=3,998, lactate resulted N=3,994, otherwise N=3,999. Data detail in Tables 3, 6, and 9.

### Hours in the intensive care unit

For patients discharged alive who went to the ICU, treatment with IV fluids within 3 hours after sepsis onset was associated with the largest estimated reduction in average hours spent in the ICU (25.93 hours for the full cohort, 35.06 hours for severe sepsis, and 41.99 hours for septic shock) (Table 4). The next most impactful treatment for the full cohort was IV fluid initiation within 1 hour (20.83-hour reduction); for severe sepsis and septic shock, IV fluids and broad-spectrum antibiotics within 3 hours of sepsis onset was estimated to have the next-largest impact (21.48-hour reduction and 23.5-hour reduction, respectively). For all three cohorts, treatment with IV fluids and antibiotics together within 3 hours of sepsis onset (delivered 69.5% of the time) was the best opportunity for change to current practice, with an estimated intervention effect of 4.85 fewer ICU hours overall, 5.07 for severe sepsis, and 3.85 for septic shock.

When measured from ED arrival (Table 7), IV fluid initiation within 3 hours was still associated with the largest estimated reductions in average ICU hours (23.78 full cohort, 19.06 severe sepsis, 35.66 septic shock). The largest estimated impact of a change to current practice for the full cohort was combined initiation of IV fluids and antibiotics within 3 hours (4.18 fewer hours), for severe sepsis was IV fluid initiation within 1 hour (5.74 fewer hours), and for septic shock was IV fluid initiation within 3 hours (3.36 fewer hours).

As with mortality and hospital LOS, measurement of vital signs within 10 minutes of ED arrival was not found to have an impact on hours spent in the ICU (Table 10). Lactate measurement within 3 hours was associated with an estimated 20.15 fewer ICU hours overall (19.99 severe sepsis, 13.34 septic shock). Lactate measurement within one hour was actually associated with an increase in ICU hours, which is most likely an indication that the 37.1% of admissions that had a lactate resulted very quickly were for sicker individuals. Sepsis screening within 15 and 30 minutes had an estimated impact of 6.04 and 6.93 fewer ICU hours in the full cohort, but none within those with severe sepsis or septic shock. The estimated impact of any change to current practice was small – if the percentage of patients with a lactate resulted within 3 hours of ED arrival increased from 83.2% to 100%, we could see a reduction of 3.49 average hours spent in the ICU (2.99 for severe sepsis, no change for septic shock). Figures 14-19 display the forest plots for Tables 4, 7, and 10.

**Figure 14.**
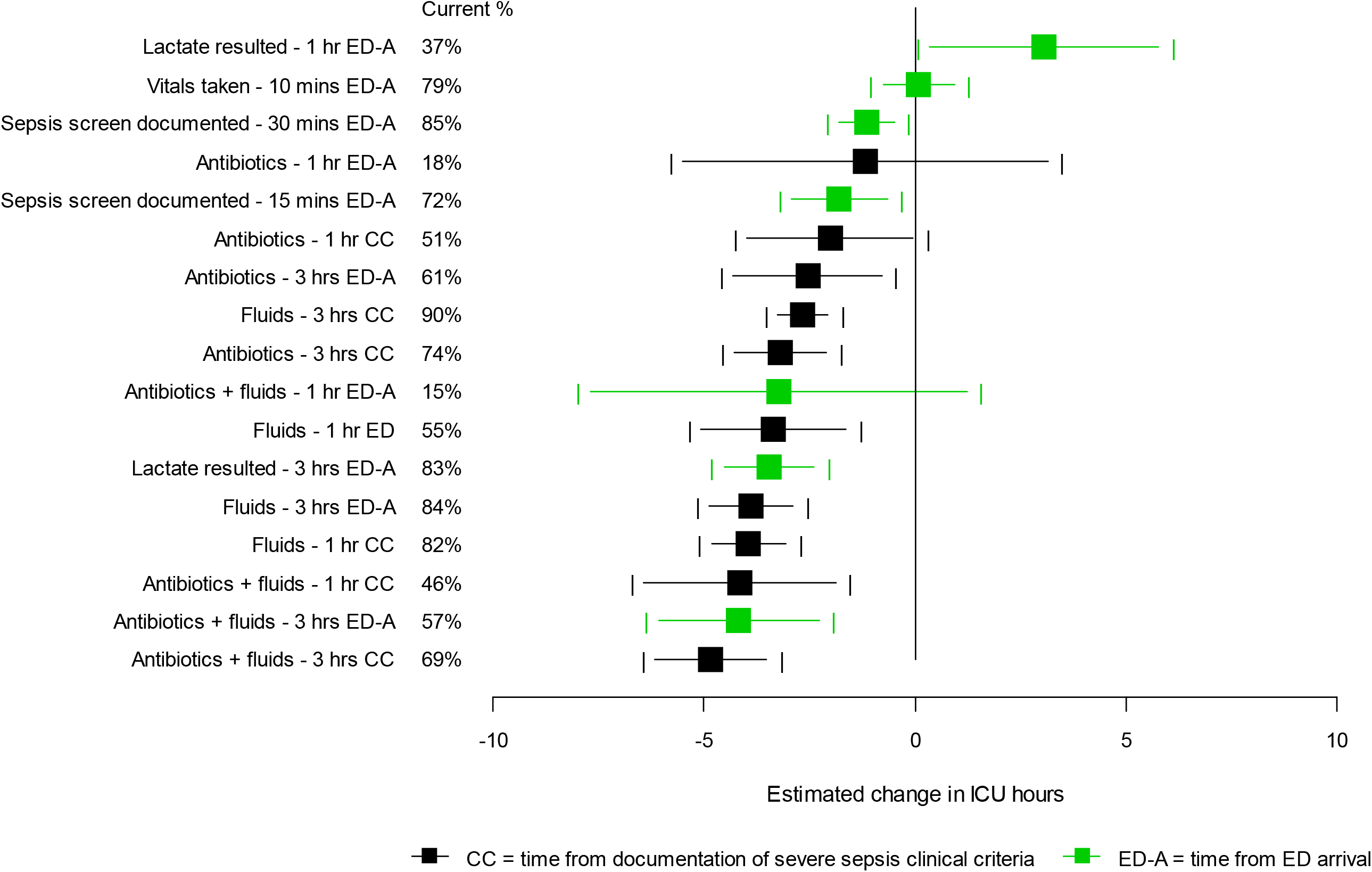
Estimated opportunity to impact hours spent in the ICU for patients discharged alive (100% success vs. current %). Point estimates and 95% confidence intervals are shown. The vertical line at zero indicates no impact. Full cohort; sepsis screen/vital signs N=10,543, lactate resulted N=10,113, otherwise N=10,544. Data detail in Tables 4, 7, and 10.

**Figure 15.**
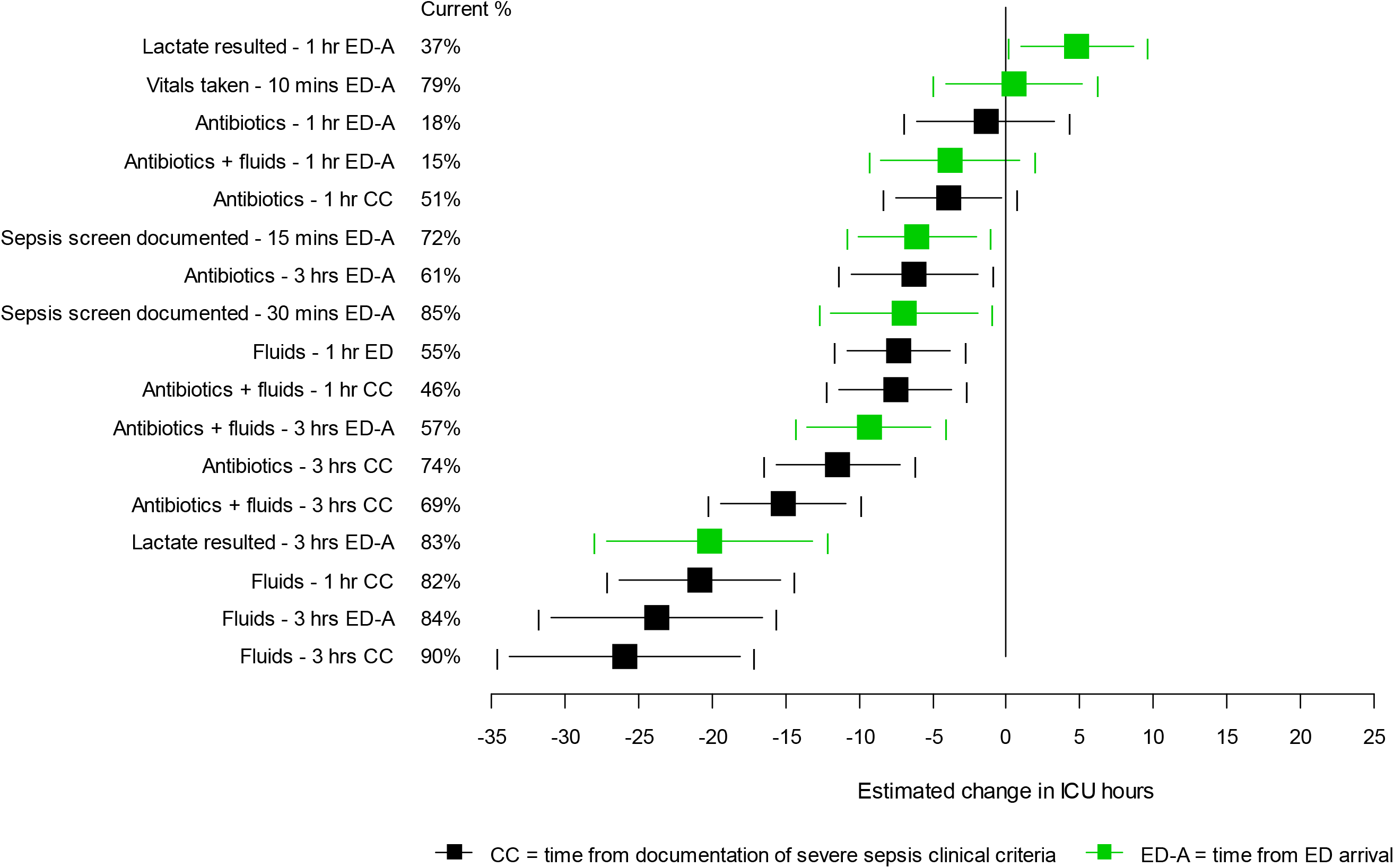
Estimated impact of timely identification and treatment on hours spent in the ICU for patients discharged alive (100% success vs. 0% success). Point estimates and 95% confidence intervals are shown. The vertical line at zero indicates no impact. Full cohort; sepsis screen/vital signs N=10,543, lactate resulted N=10,113, otherwise N=10,544. Data detail in Tables 4, 7, and 10.

**Figure 16.**
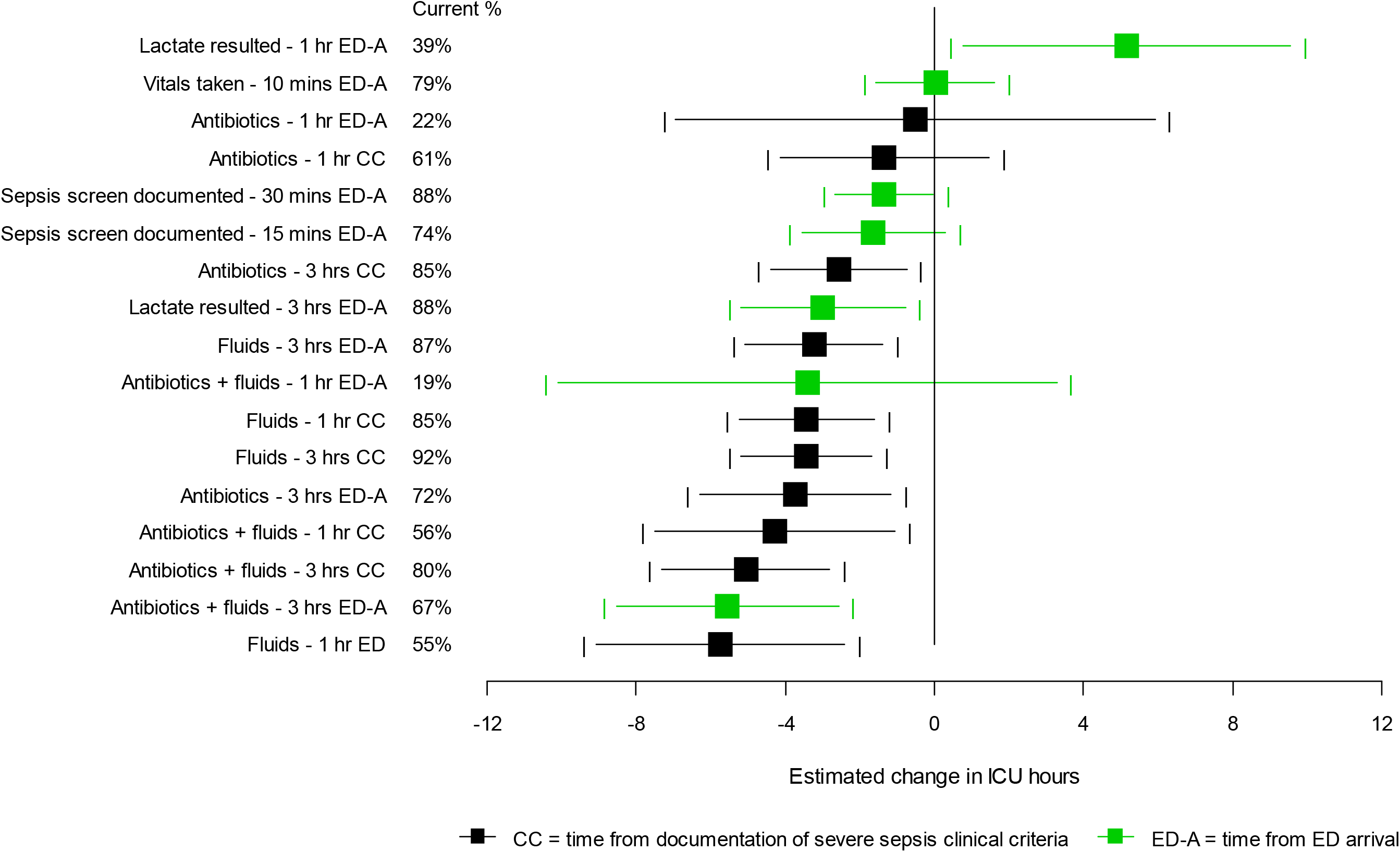
Estimated opportunity to impact hours spent in the ICU for patients discharged alive (100% success vs. current %). Point estimates and 95% confidence intervals are shown. The vertical line at zero indicates no impact. Severe sepsis cohort; lactate resulted N=2,390, otherwise N=2,395. Data detail in Tables 4, 7, and 10.

**Figure 17.**
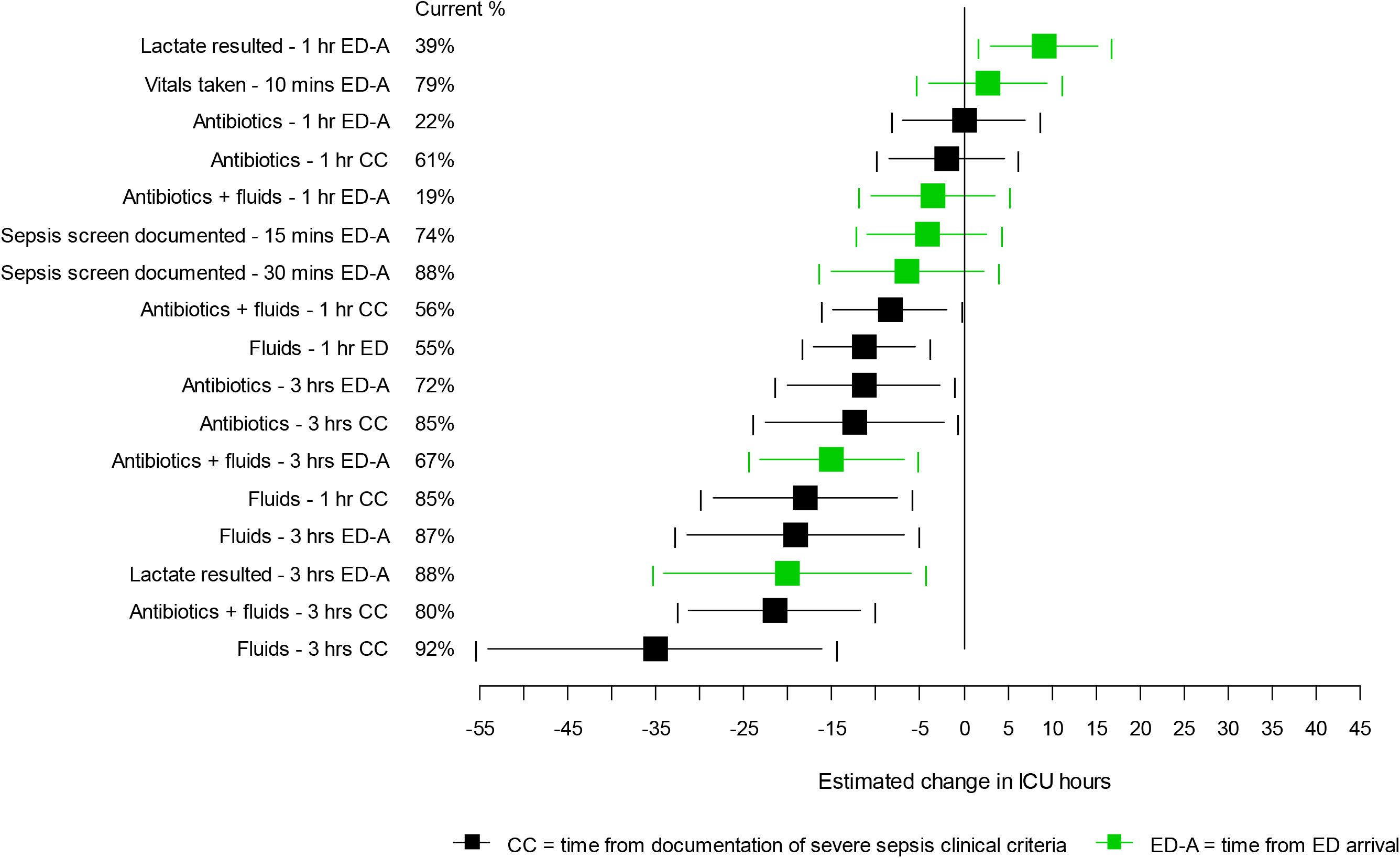
Estimated impact of timely identification and treatment on hours spent in the ICU for patients discharged alive (100% success vs. 0% success). Point estimates and 95% confidence intervals are shown. The vertical line at zero indicates no impact. Severe sepsis cohort; lactate resulted N=2,390, otherwise N=2,395. Data detail in Tables 4, 7, and 10.

**Figure 18.**
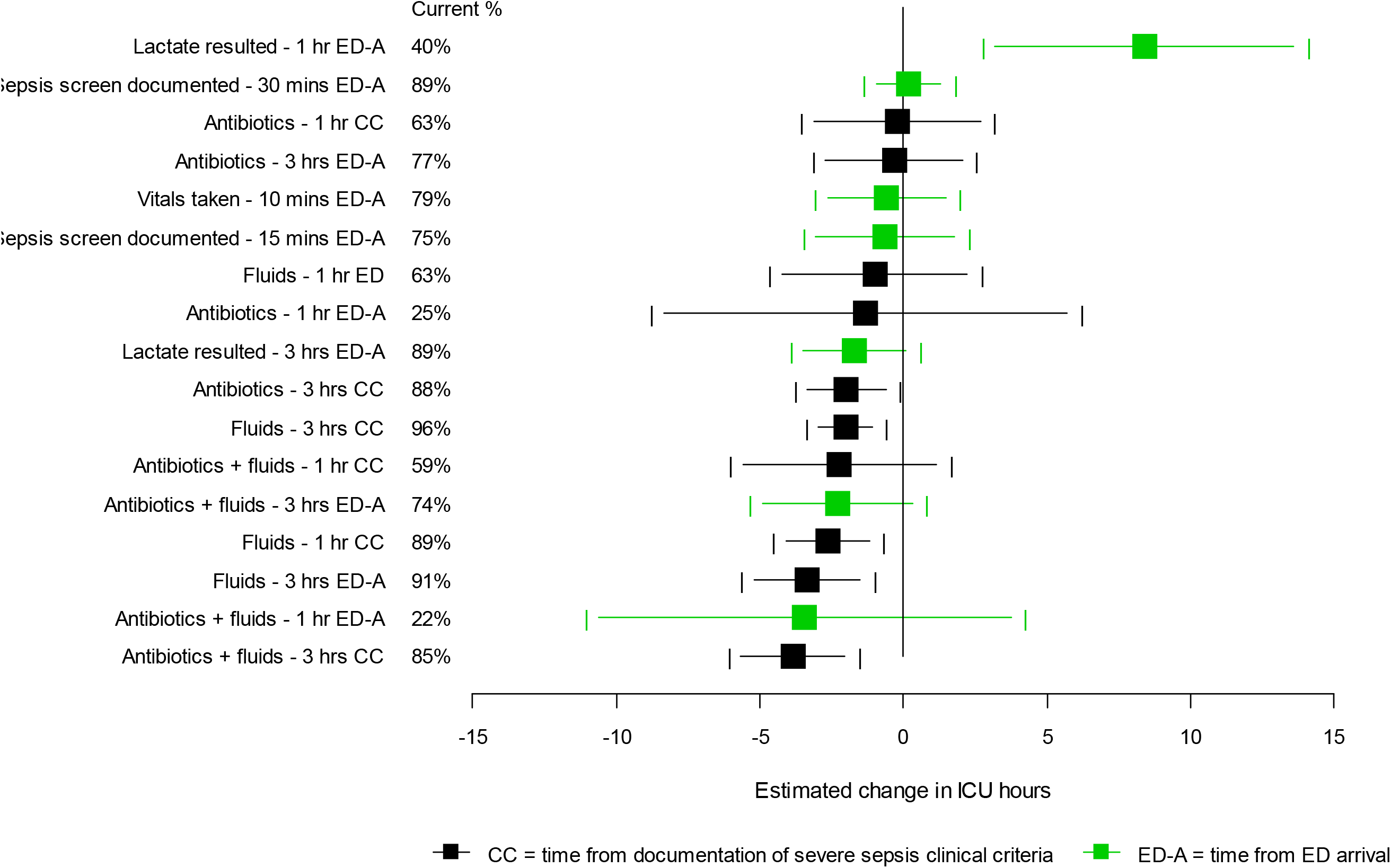
Estimated opportunity to impact hours spent in the ICU for patients discharged alive (100% success vs. current %). Point estimates and 95% confidence intervals are shown. The vertical line at zero indicates no impact. Septic shock cohort; sepsis screen/vital signs N=3,487, lactate resulted N=3,484, otherwise N=3,488. Data detail in Tables 4, 7, and 10.

**Figure 19.**
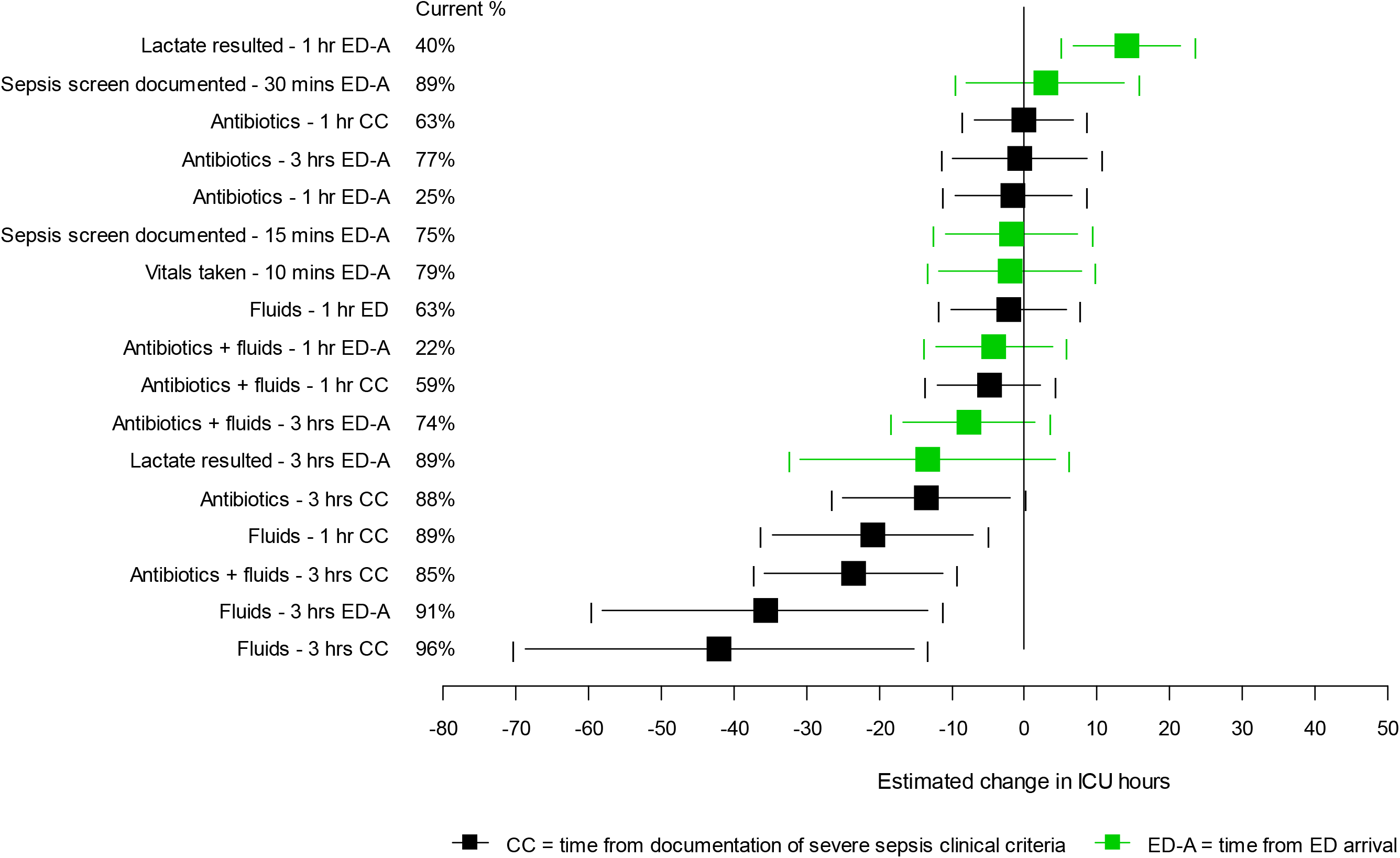
Estimated impact of timely identification and treatment on hours spent in the ICU for patients discharged alive (100% success vs. 0% success). Point estimates and 95% confidence intervals are shown. The vertical line at zero indicates no impact. Septic shock cohort; sepsis screen/vital signs N=3,487, lactate resulted N=3,484, otherwise N=3,488. Data detail in Tables 4, 7, and 10.

### Study Limitations

This study has several limitations. First, prospective identification of sepsis cases is challenging – as highlighted by the fact that 45.1% of admissions in the study population did not go on to have a sepsis discharge diagnosis. ED clinicians, however, may have to initiate treatment before sepsis has been confirmed, so we have attempted to provide a more complete picture (than would be provided by considering those who have a sepsis discharge diagnosis only) by showing the full cohort of individuals who could have been treated, as well as looking at the subset of admissions that resulted in severe sepsis and septic shock diagnoses. Second, in attempting to include measures of severity of condition at ED arrival, it was necessary to use time to sepsis onset (according to clinical criteria as described in this report) and the presence or absence of the clinical criteria at sepsis onset as proxies for severity at presentation – even though these values were in reality measured after ED arrival. For estimates that measure time from the door, this could potentially create complications with the time-ordering of events. Given that it is not unreasonable to assume that measured lab values and vital signs simply revealed issues that were present upon ED arrival, this was determined to be an acceptable risk for this analysis, and excluding measures of severity at presentation would have negatively impacted any ability to model the outcomes effectively. Finally, the structure of the analysis does not allow for comparison *between* treatments – for example, to answer the question of what *additional* impact treatment within 1 hour could have over treatment within 3 hours. This could be easily addressed in a future analysis with these data, but was not the focus of this particular study. Given that the focus of this study was on the timing of treatment initiation, the volume of IV fluids given during sepsis treatment was not considered. This is another area of potential future study.

## CONCLUSIONS

This analysis suggests that treatment of patients with IV fluids and broad-spectrum antibiotics earlier had a positive impact on patient outcomes at Sutter Health during 2016-17. Prompt sepsis screening appeared to also be making a difference. The greatest opportunity presented, in terms of potential impact if current practice could be altered, was in the area of combined initiation of IV fluids and broad-spectrum antibiotics within one hour of sepsis presentation.

## Data Availability

Data are not currently available for external use.

## APPENDIX

**Figure A.**
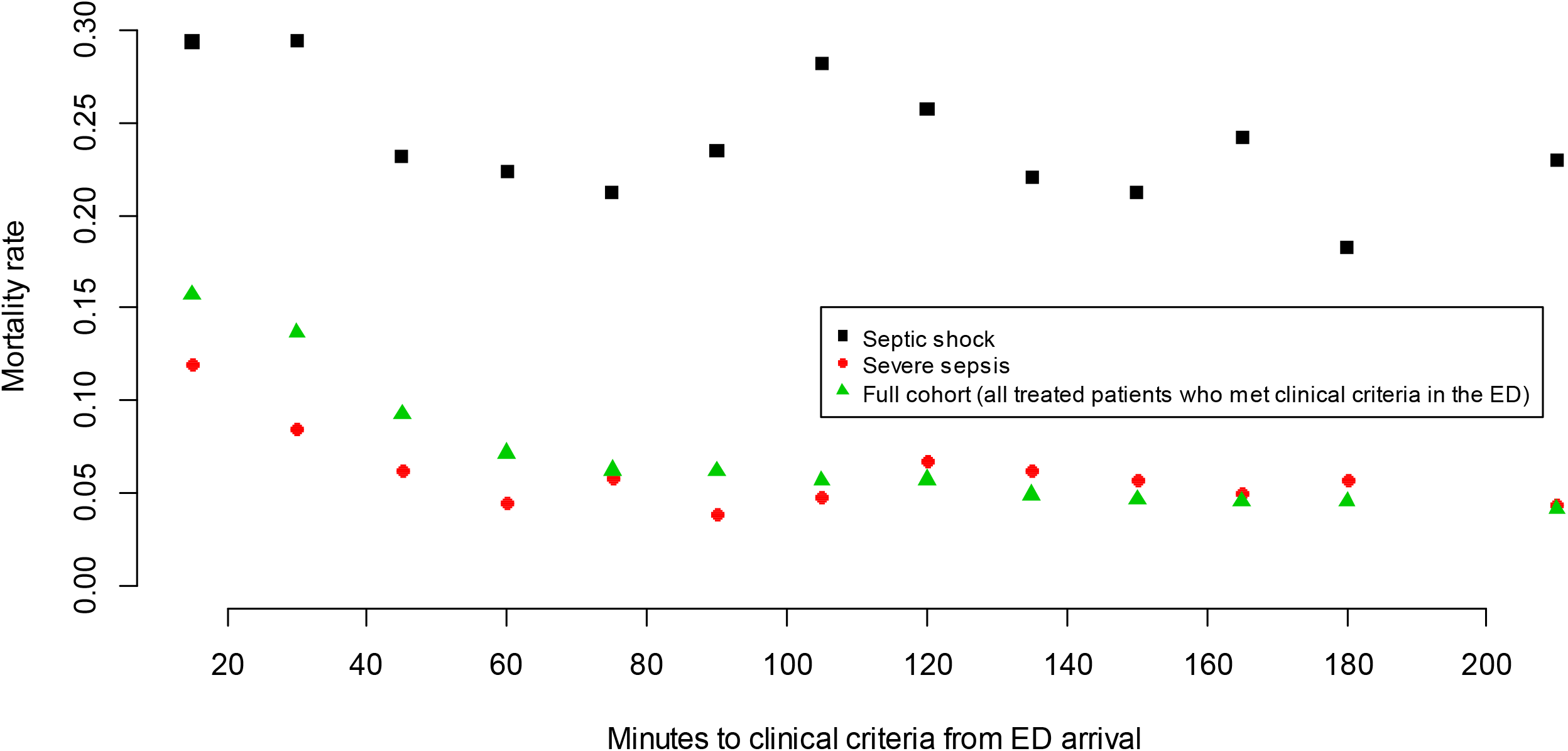
Relationship between time to first appearance of severe sepsis clinical criteria in the medical record and mortality

## Notes

### Competing Interest Statement

The authors have declared no competing interest.

### Funding Statement

No external funding was received.

### Author Declarations

Sutter Health IRB

